# Strong Protein Responses to Acute Infection Control General Post-Viral Sequelae and SARS-CoV-2-specific Neurocognitive Sequelae

**DOI:** 10.64898/2026.08.27.26361488

**Authors:** Theodore G. Liou, Ryan J. Andrews, Heather S. Battey, Brenda L. Bass, Luis G. Vargas Buonfiglio, Barbara C. Cahill, James E. Cox, Scott Gibson, Stephen C. Hartsell, Nathan Hatton, Mark W. Hazel, My N. Helms, Judy L. Jensen, Christiana Kartsonaki, Jesse Kupfer, Yanping Li, Filipa Blasco Tavares Pereira Lopes, Allison Manuel, Marco Marchetti, James E. Marvin, Elizabeth A. Middleton, Patrice Mimche, Kristyn A. Packer, Robert Paine, Rhonda D. Szczesniak, Anne B. Sturrock, Anwar Tandar, E. Bart Tarbet, Andrew Ulrich, Derek Warner, Kristi Jo Warren, Allison M. Weis, Elizabeth Zimmerman, Sarang K. Yoon, Micah G. Ownbey, Scott T. Youngquist, Frederick R. Adler

**Affiliations:** Division of Respiratory, Critical Care and Occupational Pulmonary Medicine, Department of Internal Medicine, Spencer Eccles Fox School of Medicine, University of Utah, 26 North Mario Capecchi Drive, Salt Lake City, UT 84112, USA; Department of Pediatrics and Department of Population Health Sciences, Spencer Eccles Fox School of Medicine, University of Utah, Salt Lake City, UT, USA and Department of Biological and Irrigation Engineering, Utah State University, Logan, UT, USA; Center for Quantitative Biology, University of Utah, Salt Lake City, UT, USA; Department of Biochemistry, Spencer Eccles Fox School of Medicine, University of Utah, Salt Lake City, UT, USA; Department of Mathematics, Imperial College London, London, UK; Metabolomics, Proteomics and Mass Spectrometry Core, Spencer Eccles Fox School of Medicine, University of Utah, Salt Lake City, UT, USA; The Institute for Antiviral Research, College of Veterinary Medicine, Logan, UT 84322, USA; Department of Emergency Medicine, Spencer Eccles Fox School of Medicine, University of Utah, Salt Lake City, UT, USA; Division of Gastroenterology, Department of Internal Medicine, Spencer Eccles Fox School of Medicine, University of Utah, Salt Lake City, UT, USA; Clinical Trial Service Unit & Epidemiological Studies Unit and Medical Research Council Population Health Research Unit, Nuffield Department of Population Health, University of Oxford, Oxford, UK; DNA Sequencing Core Laboratory, University of Utah, Salt Lake City, Utah, USA; Flow Cytometry Core Laboratory, University of Utah, Salt Lake City, Utah, USA; Department of Pathology, University of Utah, Salt Lake City, Utah, USA; current appointments Departments of Dermatology and of Medicine, Indiana University, Indianapolis, Indiana, USA; Department of Veterans Affairs Medical Center, Salt Lake City, UT, USA; Department of Pediatrics, Divisions of Biostatistics & Epidemiology and Pulmonary Medicine, Cincinnati Children’s Hospital and University of Cincinnati, Cincinnati, Ohio, USA; Division of Cardiology, Department of Internal Medicine, Spencer Eccles Fox School of Medicine, University of Utah, Salt Lake City, UT, USA; Division of Occupational and Environmental Health, Spencer Eccles Fox School of Medicine, Rocky Mountain Center for Occupational and Environmental Health, Salt Lake City, Utah, USA; Department of Mathematics and School of Biological Sciences, University of Utah, Salt Lake City, UT 84112, USA

**Author notes:** These authors shared roles as Senior Authors for Clinical Work (Micah G. Ownbey, Scott T. Youngquist). Senior Author for Analytical Work (Frederick R. Adler).

**Keywords:** Post-Acute Infection Syndrome, Respiratory Viral Infection, Viral Syndrome, SARS-CoV-2, COVID19, Post-Acute Sequelae of COVID19, Long COVID, Innate Immunity, Editosome, Proteomics

## Abstract

After serious infections, many survivors fully recover while others face persistent symptoms including neurocognitive dysfunction, cardiopulmonary instabilities and disabling fatigue, classified broadly as Post-Acute Infection Syndromes (PAIS). Post-Acute Sequelae of COVID19 (PASC) is one special case. To test whether acute infection initiates PAIS, we analyzed proteins in 173 saliva samples collected during severe respiratory viral infections and tracked patient outcomes for five years. We discovered 44 proteins, all derived from interferon stimulated genes, that respond strongly to acute infection. Rates of post-acute syndromes were similar regardless of infection, but the types of Sequelae differed by virus. High expression of 19 proteins predict low General Sequelae rates across all viruses. High expression of 23 overlapping proteins predict low Neurocognitive Sequelae rates only with SARS-CoV-2 infections. We propose that low expressions of these proteins, which control viral replication, innate immunity, and secondary autoimmunity, combine to precipitate PAIS and PASC.

**Highlights:**

- Acute viral infections elicit interferon stimulated gene (ISG) protein responses.
- 23 ISG proteins prevent post-infection General Sequelae for all viral infections.
- 19 ISG proteins prevent post-infection Neurocognitive Sequelae only with SARS-CoV-2.
- Poor protein responses induce distinct post-viral syndromes with or without COVID19.

## Introduction

Acute infections affect all human communities. Infections may be severe, leading to hospitalization and intensive care when fever, shortness of breath, confusion, malaise and many other symptoms indicate physiologic deterioration that threatens survival. Depending on the infection, different proportions of people rapidly and fully recover while others die from organ failures potentially magnified by pre-existing comorbidities.^1^ A portion of those with acute illness, including some with mild disease, transition to debilitating post-acute states that are broadly characterized as post-acute infection syndromes (PAIS).^2,3^

Respiratory viruses can rapidly infect entire communities by airborne spread and cause a pandemic. The SARS-CoV-2 pandemic, which caused more than 7 million deaths January 2020 to December 2025,^4^ focused attention on one special case of PAIS, Post-Acute Sequelae of COVID19 (PASC). PASC accounted for 211 million disability-adjusted life-years lost in 2021, surpassing all other causes.^5^ Disability persists months to years and currently affects millions in the US and tens to hundreds of millions worldwide. Estimated global economic losses from PASC exceed 1 trillion US dollars annually.^6^

Also known as Long COVID, PASC follows 10-25% of acute infections with illnesses lasting three or more months to years.^7^ Severe symptoms impair work, school and activities of daily living^3,8^ and include neurocognitive dysfunction, chronic fatigue, cardio-pulmonary instabilities and post-exertional malaise, among many other symptoms.^9,10^ The symptoms may be classified into three broad categories: **Neurocognitive Sequelae** including brain fog, anxiety and headaches; **General Sequelae** including fatigue, sleep disturbances and musculoskeletal complaints; and **Respiratory Sequelae** including chronic shortness of breath, cough and hypoxemia.

Multiple investigations have identified many potential underlying mechanisms,^8,11^ yet PASC still lacks diagnostic clarity and specific treatments.^12^ PASC shares with PAIS the lack of an explanatory pathobiology that links acute disease to chronic sequelae. Because some findings associated with PASC appear during the acute phase of COVID19,^13,14^ we hypothesized that acute infection can initiate a post-viral syndrome, more specifically, that acute SARS-CoV-2 respiratory infection induces a virus-specific human protein signature detectable in saliva that anticipates PASC. Because PASC is a special case of PAIS, we surmised that acute protein expression signatures might provide insights into PAIS more broadly.

To answer the question whether acute infection initiates a post-viral syndrome, we conducted a prospective observational study during the SARS-CoV-2 pandemic of people seeking emergency care for acute respiratory viral syndromes. We collected saliva samples and analyzed protein responses to identify acute signatures that might initiate post-viral mechanisms. To link acute protein expression with PASC manifestations, we counted post-acute viral symptoms,^15^ and rigorously analyzed the data to determine whether different acute protein expression levels preceded development of three post-viral symptom classes: **Neurocognitive, General** and **Respiratory Sequelae**.

We found that many proteins respond vigorously to acute viral infections. These proteins derive from interferon stimulated genes^16^ and include proteins that directly oppose viral replication and control innate immune and autoimmune responses triggered by double-stranded RNA.^17^ Insufficient acute expression of these proteins was strongly linked to persistent General Sequelae across all viruses and Neurocognitive Sequelae specific to SARS-CoV-2.

## Results

### Clinical Data Acquisition

#### Patient Recruitment

After institutional review board approval (IRB_00134424), we prospectively recruited 236 University of Utah Emergency Department (ED) patients between January 2021 and May 2023 (**Figure S1**) after written informed consent. Patients presented with respiratory viral symptoms, after urgent referral from other clinical settings for testing and care, or with worsening symptoms after an initial assessment. We excluded 23 patients unable to produce saliva and 40 patients with more than 30 days of symptoms before enrollment. We restricted analysis to the remaining 173 (**Table 1, Figure S2**).

**Table 1.** Patient Characteristics.

| Characteristic | Measurements | Patients with Data |
| --- | --- | --- |
| ***Demographics*** |  |  |
| Years of Age* | 49.46 (SD = 18.85) | 173 |
| Female Sex† | 87 (0.50) | 173 |
| White Race† | 122 (0.70) | 173 |
| Number of People in Home* | 3.21 (SD = 2.02) | 158 |
| Vaccinated at least once† | 58 (0.335) | 173 |
| Days from Vaccination to ED Visit | 24.5 (0 to 587) | 56 |
| Vaccination Dates | 6 Jan 2021 to 8 Dec 2022 | 56 |
| Emergency Department Visit Dates | 11 Jan 2021 to 9 May 2023 | 173 |
| Prior SARS-CoV-2 Infection Dates | 1 Mar 2020 to 4 Jan 2023 | 71 |
| ***Emergency Department Findings and Disposition*** |  |  |
| SARS-CoV-2 Infection Detected† | 100 (0.58) | 173 |
| Days from first symptom* | 8.02 (SD = 6.99) | 173 |
| Hypoxemia† | 48 (0.28) | 173 |
| Oxygen Percent Saturation* | 92.66 (SD = 5.81) | 172 |
| Respiratory Rate* | 19.96 (SD = 4.76) | 171 |
| Hospitalization† | 72 (0.42) | 173 |
| Discharged to Home† | 101 (0.58) | 173 |
| Discharged to Home on Oxygen† | 7 (0.07) | 99 |
| ***Past Medical Conditions*** |  |  |
| Prior SARS-CoV-2 Infection† | 23 (0.133) | 173 |
| Chronic Infections† | 15 (0.087) | 173 |
| Central Nervous System Disease† | 46 (0.266) | 173 |
| Mental Health Disease† | 79 (0.457) | 173 |
| Hypertension† | 65 (0.376) | 173 |
| Diabetes mellitus† | 27 (0.156) | 173 |
| Other Endocrinopathies† | 23 (0.133) | 173 |
| Obesity† | 41 (0.237) | 173 |
| Heart Disease† | 66 (0.382) | 173 |
| Lung Disease† | 73 (0.422) | 173 |
| Gastrointestinal Disease† | 65 (0.376) | 173 |
| Hepatitis† | 9 (0.052) | 173 |
| Rheumatologic Diseases† | 54 (0.312) | 173 |
| Cancer† | 30 (0.173) | 173 |
| Clotting Issues or Bleeding Diatheses† | 15 (0.087) | 173 |
| Renal Disease or Hemodialysis† | 26 (0.15) | 173 |
| Solid Organ Transplantation† | 11 (0.064) | 173 |
| ***Habits*** |  |  |
| Substance Abuse† | 25 (0.145) | 173 |
| Cigarette Smoking† | 24 (0.139) | 173 |
| Smoking or Vaping Any Substance† | 47 (0.272) | 173 |
| ***Health Care Usage in Prior Year*** |  |  |
| Clinic§ | 1266 (7.318) | 173 |
| ED§ | 195 (1.127) | 173 |
| Hospitalization§ | 124 (0.717) | 173 |
| ***Health Care Usage in Post Enrollment Year*** |  |  |
| Clinic§ | 1253 (7.243) | 173 |
| ED§ | 262 (1.514) | 173 |
| Hospitalization§ | 76 (0.439) | 173 |

\* Mean (Standard Deviation)
† Number of Patients (Decimal Fraction of Patients with Data)
§ Total Visit Counts (per patient mean)
¶ Count obscured to protect patient health information

**Table 1. Patient Characteristics**
| Characteristic | Measurements | Patients with Data |
| --- | --- | --- |
| ***Follow-up*** |  |  |
| Last Contact Dates (Includes Deaths) | 19 Jan 2021 to 19 Nov 2025 | 173 |
| Patients with Any Post-Viral Sequelae<sup>†</sup> | 90 (0.52) | 173 |
| Total Sequelae (Sequelae per patient) | 389 (2.25) | 173 |
| Death<sup>†</sup> | 27 (0.16) | 173 |
| ***Housing Information*** |  |  |
| Homes Judged Too Small<sup>†</sup> | 16 (0.107) | 149 |
| Homes Judged Right Sized<sup>†</sup> | 119 (0.799) | 149 |
| Homes Judged Too Large<sup>†</sup> | 14 (0.094) | 149 |
| Apartment, Condominium, House<sup>†</sup> | 154 (0.911) | 169 |
| Other Living Situations<sup>†</sup> | 7 (0.042) | 169 |
| Unhoused<sup>†</sup> | 8 (0.047) | 169 |
| ***Sample Characteristics*** |  |  |
| Milliliters of Volume<sup>*</sup> | 1.89 (SD = 1.07) | 173 |
| Protein, mg/sample<sup>*</sup> | 3.37 (SD = 1.79) | 173 |
| RNA, µg/sample<sup>*</sup> | 3.27 (SD = 2.50) | 123 |
| Number of Poor Quality Saliva Samples<sup>‡</sup> | <5 | 173 |

<sup>\*</sup> Mean (Standard Deviation)
<sup>†</sup> Number of Patients (Decimal Fraction of Patients with Data)
<sup>§</sup> Total Visit Counts (per patient mean)
<sup>‡</sup> Count obscured to protect patient health information

#### ED Presentation

Participants had widely ranging age and multiple comorbidities. They were roughly equally divided by sex and SARS-CoV-2 detection. Approximately one-third had prior confirmed SARS-CoV-2 infections, one-third were vaccinated, and about a quarter reported habits associated with lung or systemic disease such as smoking. Racial, ethnic and housing information suggested representation spanning social and economic strata. Nearly half of the study patients were either hospitalized or discharged home with newly prescribed supplemental oxygen, and 16% died within 5 years.

#### ED Samples

We non-invasively collected saliva. No one refused the request, and 173 produced sufficient saliva for analysis. Saliva samples averaged 1.89 ml (Standard Deviation [SD] = 1.07 ml, range = 0.01 to 5.0 ml). Following centrifugation, the pellet fractions contained 3.37 mg of protein per sample on average (SD = 5.44 mg, range = 0.17 to 9.62 mg). Pellets from 134 of the participants had 3,267 ng of RNA per sample on average (SD = 2,497 ng, range = 32.9 to 10,396.4 ng).

#### Counting Sequelae after ED Presentation

During 5 years of follow up, 90 patients reported 389 persistent post-viral symptoms or 2.25 sequelae per overall study participant, a burden similar to prior reports.^15^ We categorized symptoms as Neurocognitive (78), General (187) and Respiratory (124) Sequelae (**Table S1**). Sequelae occurred at similar rates following SARS-CoV-2 (2.23 sequelae per participant) or other respiratory viral syndromes (2.27 sequelae per participant). Similar numbers of individuals with or without SARS-CoV-2 infection experienced sequelae (χ-square = 1.18, *p* = 0.28).

### Laboratory Data Acquisition

#### Proteomics

We performed quantitative proteomics on 120 ng protein samples using trapped ion mobility spectrometry coupled to time of flight mass spectrometry (**timsTOF-MS**). We measured spectral intensities for 6,561 human proteins that we identified by finding at least one unique peptide per protein. We considered measurements below the level of detection as partially informative and values within detection limits as fully informative. We excluded 238 proteins because their measurements were fully informative for fewer than 30 participants, leaving 6,323 proteins.

Plotting spectral intensities according to sample day after first symptom, we observed that many anti-viral^18–22^ proteins increased and decreased during illness **(Figure 1A and 1B)**. By focusing on this clinically relevant behavior to reduce the number of proteins for analysis and using a different scientific question than in subsequent analysis phases, we avoided post-selection inference issues.^23^ We retained proteins for further study based on a covariate-adjusted quadratic regression of each spectral intensity against time since first reported symptom.^24^ (See **Methods** *Data Exploration and Quadratic Regression* for details).

**Figure 1.**
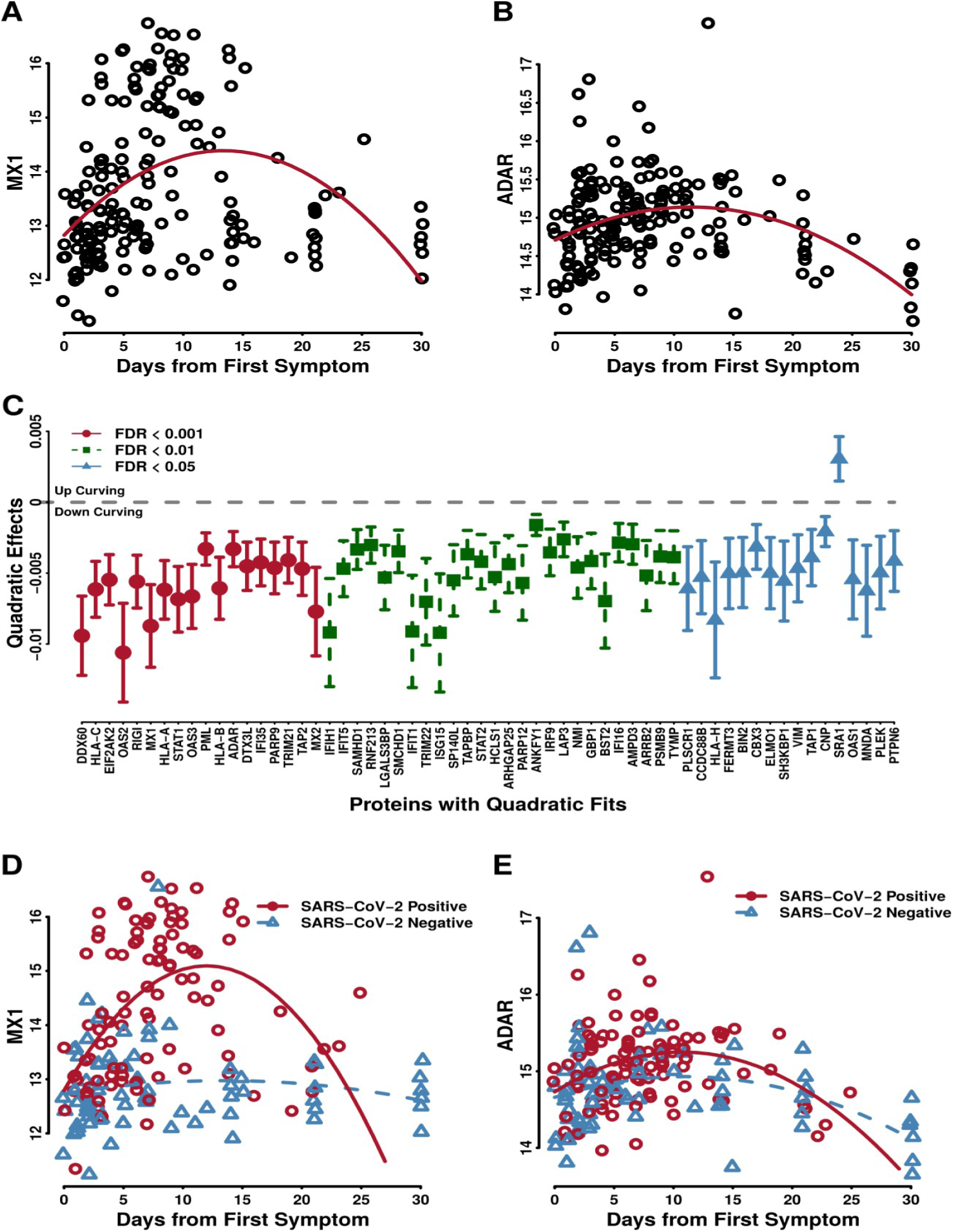
Proteins Associated with Clinical Course of Viral Syndromes. We evaluated proteins for quadratic relationships with the number of days from first reported symptom. **(A) MX1 Spectral Intensity against Days from First Symptom.** Many patients have increased spectral intensities of MX1 protein (Uniprot Designation P20591) while others do not. Quadratic regression of MX1 with days from first symptom was highly significant (*p* = 2.28×10^-08^). **(B) ADAR Spectral Intensity against Days from First Symptom.** Spectral intensities of ADAR protein (Uniprot P55265) show less variation at specific time points, and significant quadratic model fit was highly significant (p = 4.99×10^-07^). **(C) Systematic Evaluation of All Detected Proteins by quadratic regression of days from first symptom.** One hundred and one proteins had quadratic fits with False Discovery Rate-adjusted p < 0.05. We regarded the 44 proteins with *p_FDR_* < 0.01 as clinically relevant. Points ranked by *p*-value show the quadratic effects (curvature) with 95% confidence intervals for the 80 most significant proteins. **(D) MX1 and SARS-CoV-2.** MX1 measurements plotted against time in SARS-CoV-2 positive patients had significant curvature (*p* = 6.88×10^-07^) contrasting sharply with values derived from patients with other viral syndromes (*p* = 0.29). **(E) ADAR and Viral Syndromes.** Some proteins followed a different pattern over time as illustrated by ADAR. ADAR spectral intensities significantly rose and fell after symptoms started with similar degrees of curvature for SARS-CoV-2 positive patients (*p* = 0.001) and all others with viral syndromes (*p* = 0.008).

#### Quadratic Regression Analyses of Protein Spectral Intensity by Time Since First Symptom

We found 828 proteins with significant quadratic terms (*p* < 0.05) meaning that they significantly rose and fell (downward-curving) or fell and rose (upward curving) during the course of acute disease. Of these, 101 proteins had False Discovery Rate-(FDR)^25^-adjusted *p-*value (***p*_FDR_**) < 0.05 (**Table S2**), and 44 had *p*_FDR_ < 0.01. We focused further analyses on these 44 proteins, which all had downward-curving quadratic fits (**Figure 1C**), and seemed to have strong clinically relevant responses to infection. All 44 derived from interferon stimulated genes (**ISG**s).^26^ Further exploration^27^ demonstrated that some proteins, for example, MX1, responded over time to SARS-CoV-2 infection but not other viral syndromes **(Figure 1D**). In contrast, other proteins, for example, ADAR, had similar patterns for all pathogens **(Figure 1E**).

#### Multiplex Protein Assays

We measured water-soluble inflammatory biomarkers important in human viral infections from saliva supernatants in 169 of the 173 patients using a multiplex enzyme-linked immunosorbent assay (**ELISA**). All proteins detected had fully informative measurements for more than 30 patients (range: 54 to 167); none of these proteins were also detected by mass spectrometry (**MS**). We found no significant quadratic terms in regression against days from first symptom (unadjusted *p <* 0.05, **Table S3**).

#### RNA-sequencing to Identify Additional Viral Pathogens

We performed bulk next-generation RNA-sequencing (**RNA-Seq**) to identify additional viral agents (**Table S4, Top**). Using Kraken2 sequence processing,^28^ we identified viral pathogens in 54 (40%) of 134 patients with sufficient RNA for sequencing. We identified co-infections with two or three viruses in 14 patients including 6 with SARS-CoV-2.

Sixty-eight patients had both PCR and RNA-Seq assays using samples obtained within minutes of each other. In this group, PCR detected 27 SARS-CoV-2 positive patients (40%), while RNA-Seq detected 29 positives (43%, 19 overlapping) (**Table S4, Bottom**). Because the assays had similar sensitivities, we regarded samples from the entire study cohort that were positive by either or both methods as diagnostic of SARS-CoV-2 infection for analyses.

#### Avoiding Bias

We explored the potential of patient characteristics to distort the apparent relationship between acute-phase salivary proteins and post-viral sequelae. To evaluate the effects of pre-study disease burden, we applied principal components analysis^29^ to intrinsic factors, such as age and sex, and comorbidities for each patient (**Table 1**). We named the first principal component **MedPC1**; higher MedPC1 values indicate higher disease burdens (**Table S5**). Using MedPC1 as a regression variable, we found more frequent SARS-CoV-2 detection among patients with low chronic illness burdens (**Figure 2A**) establishing the need to adjust subsequent models exploring associations with SARS-CoV-2 to avoid disease specific bias.^30^

**Figure 2.**
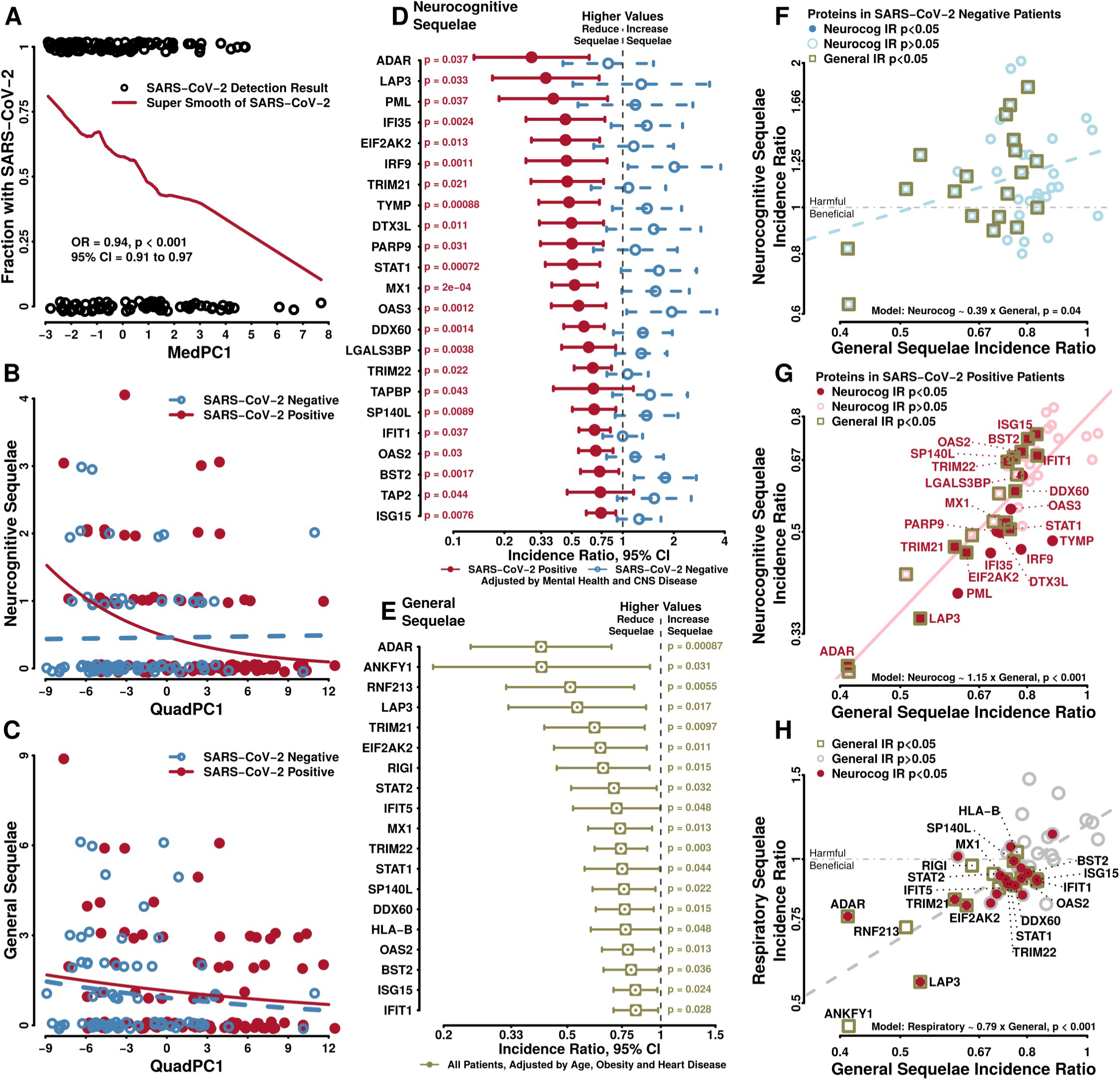
Protein Relationships with Patient Characteristics and Outcomes. **(A) Disease Specific Bias.** A super smooth plot of nucleic acid detections of SARS-CoV-2 (0 = negative, 1 = positive, jittered to show the density of patient results) by the first principal component of intrinsic and comorbidity variables (MedPC1) shows that patients with few medical conditions were highly likely to have SARS-CoV-2 infection while patients with severe chronic disease burdens were unlikely. Logistic regression of SARS-CoV-2 detection and MedPC1 (OR = 0.94, *p* < 0.001) revealed a strong disease-specific bias requiring confounder adjustments in models involving SARS-CoV-2 infection status. **(B) Neurocognitive Sequelae Counts and Clinically Relevant Proteins.** The negative binomial model of the first principal component derived from expression of 44 clinically relevant proteins (QuadPC1) as independent variable shows that neurocognitive sequelae counts are higher when QuadPC1 is low in SARS-CoV-2 positive patients (red solid curve, *p* < 0.001). Without SARS-CoV-2 infection, Neurocognitive Sequelae counts do not vary according to QuadPC1 (blue dashed line). **(C) General Sequelae Counts and QuadPC1.** General Sequelae counts are higher for lower QuadPC1. At any QuadPC1 value, SARS-CoV-2 positive individuals have more sequelae (red solid curve) than individuals without SARS-CoV-2 (blue dashed curve) demonstrating that SARS-CoV-2 infection was always worse than other viral syndromes. **(D) Incidence ratios for neurocognitive sequelae counts with 95% confidence intervals.** Red forest plots show that higher amounts of 23 proteins are associated with lower counts of Neurocognitive Sequelae in SARS-CoV-2 positive patients. SARS-CoV-2 negative patients in blue illustrate that no proteins were associated with beneficial incidence ratios; *p*-values shown are derived from full interaction models (**Table S7**). In SARS-CoV-2 positive patients, a doubling of each protein was associated with a reduction in neurocognitive sequelae by a factor equal to its incidence ratio. For example, in SARS-CoV-2 positive patients, a 1 unit increase in log_2_-transformed IRF9 (a doubling of spectral intensity) was associated with a reduction in Neurocognitive Sequelae counts by a factor of 0.25 or a 75% reduction. A 1 unit increase in log _2_-transformed ADAR was associated with a reduction in Neurocognitive Sequelae by a factor of 0.34, or a 66% reduction in counts in SARS-CoV-2 positive individuals. **(E) Incidence ratios and General Sequelae counts.** Forest plots of incidence ratios and 95% confidence intervals show that high acute expressions of 19 proteins during viral syndromes were strongly associated with protective incidence ratios for subsequent General Sequelae counts. A doubling of protein amount was associated with a reduction in General Sequelae counts by a factor equal to the incidence ratio shown. For example, a 1 unit increase in log_2_-transformed ADAR was associated with a reduction in General Sequelae counts by a factor of 0.41 or a 59% decrease. **(F) Relationship between incidence ratios for Neurocognitive and General Sequelae in SARS-CoV-2 negative patients.** In 73 individuals without SARS-CoV-2, incidence ratios of the 44 clinically relevant proteins for counts of Neurocognitive Sequelae are poorly correlated with incidence ratios for counts of General Sequelae (blue dashed line). This suggests that within a general immune response to a specific infection, only certain proteins may be associated with protective effects. **(G) Relationship between incidence ratios for Neurocognitive and General Sequelae.** In the 100 individuals with COVID19, there was excellent correlation between incidence ratios for Neurocognitive and General Sequelae counts. Different overlapping protein sets (solid dots and square boxes) are associated with significant beneficial incidence ratios for Neurocognitive and General sequelae counts. Proteins with significant incidence ratios for SARS-CoV-2 interactions are named. **(H) Relationship between Protein Incidence Ratios from Respiratory and General Sequelae count models.** After adjustment for heart and lung disease, obesity, smoking and acute hypoxemia, Respiratory Sequelae count models produced incidence ratios that were not statistically significant but were still correlated to the General Sequelae incidence ratios (coefficient = 0.53, SE = 0.08, *p* < 0.001) suggesting that Respiratory Sequelae models lacked power. Proteins with significant incidence ratios for General Sequelae counts are labeled.

We used logistic regression^31^ models to control for potential bias arising from individual clinical variables. We found mental health and central nervous system disease to be relevant adjustment variables for persistent Neurocognitive Sequelae models, and age, heart disease and obesity to be relevant for General Sequelae models. We selected lung and heart disease, obesity, cigarette smoking and acute hypoxemia in the ED as relevant adjustments for Respiratory Sequelae models. Inclusion of additional acute severity measures in any of these models failed to significantly improve model fits.

### Protein Discoveries

#### Integrated Protein Responses

We fitted negative binomial regression models^32^ of Neurocognitive, General and Respiratory Sequelae counts as dependent variables. We used SARS-CoV-2 infection status and the first principal component derived from the 44 clinically responsive proteins for each study participant as independent variables. This accomplished two goals. First we established that clinically relevant proteins for acute disease encoded information for post-acute sequelae. Second, the work established model forms for facilitating examination of single protein relationships with post-acute sequelae. We derived the first principal component from log_2_-transformed timsTOF-MS spectral intensities of the proteins which we named **QuadPC1** (**Table S6**) as the proteins were discovered using quadratic regression.

We found a strongly significant interaction between SARS-CoV-2 status and QuadPC1 for the Neurocognitive Sequelae model (**Figure 2B)**. The nature of interaction indicates that protein benefits for Neurocognitive Sequelae or that Neurocognitive Sequelae themselves follow only SARS-CoV-2 in our cohort. In the General Sequelae model, SARS-CoV-2 and QuadPC1 were independently significant and additive rather than interactive (**Figure 2C**). All other factors being equal, SARS-CoV-2 infection was associated with increased subsequent General Sequelae counts (**red curve, Figure 2C**). The additive impact of SARS-CoV-2 suggests that General Sequelae occur with equal frequency but somewhat decreased severity following non-COVID-19 infections (**blue dashed curve, Figure 2C**). We found no significant interactive or additive QuadPC1 Respiratory Sequelae models.

#### Specific Protein Responses

Twenty-three out of 44 clinically relevant proteins had significant beneficial interactions with SARS-CoV-2 positive status in negative binomial models adjusted by mental health and CNS disease for Neurocognitive Sequelae counts. The incidence ratios (**red symbols, Figure 2D**) describe the fractional decreases in Neurocognitive Sequelae associated with 1-unit increases in log_2_-transformed protein spectral intensity (**Table S7**). In contrast, no proteins had significantly protective incidence ratios in participants without SARS-CoV-2 (**blue symbols, Figure 2D**).

Nineteen out of 44 clinically relevant proteins had significant beneficial incidence ratios for General Sequelae counts in negative binomial models^32^ adjusted by age, heart disease and obesity (**Figure 2E**). The incidence ratios shown equal the fractional decrease in General Sequelae associated with a 1-unit increase in log _2_-transformed protein spectral intensity regardless of viral infection. In the presence of SARS-CoV-2 infection, associated General Sequelae counts were increased independently of protein incidence ratios (**Table S8**).

#### Correlated Protein Responses

To better understand the 44 proteins summarized by QuadPC1, we compared protein-specific incidence ratios derived from negative binomial models^32^ of Neurocognitive counts and General Sequelae. For patients with non-SARS-CoV-2 infections, the individual incidence ratios of the 44 clinically relevant proteins were weakly correlated between Neurocognitive and General Sequelae outcome models (linear regression,^32^ *p* = 0.04, **Figure 2F**). In contrast, for patients with SARS-CoV-2, individual protein incidence ratios were strongly correlated (*p* = 2.1 × 10^-14^, **Figure 2G**) suggesting that many proteins are associated with both Neurocognitive and General Sequelae outcomes. The weak results (**Figure 2F**) recall alternative explanations for significant interactions between SARS-CoV-2 and QuadPC1 in Neurocognitive Sequelae modeling by suggesting that **(1)** a substantial number of ISG-derived proteins are associated with benefit in some viral infections but not others or **(2)** Neurocognitive Sequelae occurred primarily after SARS-CoV-2 infection and rarely with other viral syndromes in our cohort.

We found no clearly significant associations between Respiratory Sequelae counts and clinically relevant proteins regardless of SARS-CoV-2 status. In **Figure 2H**, the incidence ratios for some of the 44 clinically relevant proteins identified by quadratic regression lie above the 1 value on the Y-axis, illustrating the lack of statistical significance for adjusted Respiratory Sequelae count models. However, these incidence ratios were strongly correlated with the incidence ratios observed for General Sequelae counts (*p* = 6.1 × 10^-9^). The finding suggests that our analyses may simply have lacked power for qualitatively similar individual protein responses in the Respiratory Sequelae count models.

#### Sensitivity Analyses

We limited the proteins for quadratic regression analysis to those with fully informative measurements for at least 30 patients, 6,323 proteins. Inclusion of proteins with fully informative measurements for as few as 8 patients increased the number of proteins for consideration to 6,484, resulting in finding one additional protein which had a spectral intensity against time with quadratic regression *p*_FDR_ < 0.01, but that protein had no significant relationships with post-acute sequelae counts.

Negative binomial models examining individual proteins were insensitive to adjustments with race, ethnicity, home type, occupancy or comorbidities involving fewer than 20% of participants. These comorbidities included abnormal clotting or bleeding, liver, non-diabetes endocrine, or renal diseases, active or inactive cancers, organ transplantations of any type, substance abuse, smoking, vaping, vaccination status, chronic infections, or other viral infections including other coronaviruses, enteroviruses and all other viruses combined. Models were insensitive to initial presentation findings suggestive of high severity including acute hypoxemia and requirement for inpatient admission. Models were insensitive to the number of prior COVID19 infections and time between PCR testing obtained prior to ED referral and enrollment. Leave-one-out cross-validation found no indication of outlier effects due to unusual individuals.

Sample collections began during the end of α-variant, spanned all δ-variant and extended through several ο-variant infection peaks of SARS-CoV-2 strains.^33^ Models were stable regardless of sampling dates, strongly suggesting that our results reflect stable human rather than strain-specific viral biology.

### Synthesis

#### Protein Signatures

Expression analysis using the Reactome database^34,35^ identified 15 pathways associated with Neurocognitive and General Sequelae model proteins and 5 pathways associated only with General Sequelae model proteins (**Table S9**). Ordering pathways by percentage representation by our clinically relevant proteins suggests the importance of ADAR and the editosome.^17^ Other pathways reinforce the suggestion of over-representation of interferon-related pathways in acute respiratory viral syndrome responses associated with subsequent PASC or PAIS. Pathways associated only with General Sequelae include two Respiratory Syncytial Virus-related pathways, suggesting that the proteins with beneficial incidence ratios for General Sequelae may be beneficial for other infections besides SARS-CoV-2.

#### Literature-Based Anti-Viral Mechanisms

Literature searches for the 29 proteins with beneficial relationships with Neurocognitive and General Sequelae counts identified putative anti-viral mechanisms for each (**Table 2**). Organizing these mechanisms in reference to the SARS-CoV-2 life cycle suggests that the anti-viral protein responses that we identified affect nearly every viral replication stage (**Figure 3**).

**Figure 3.**
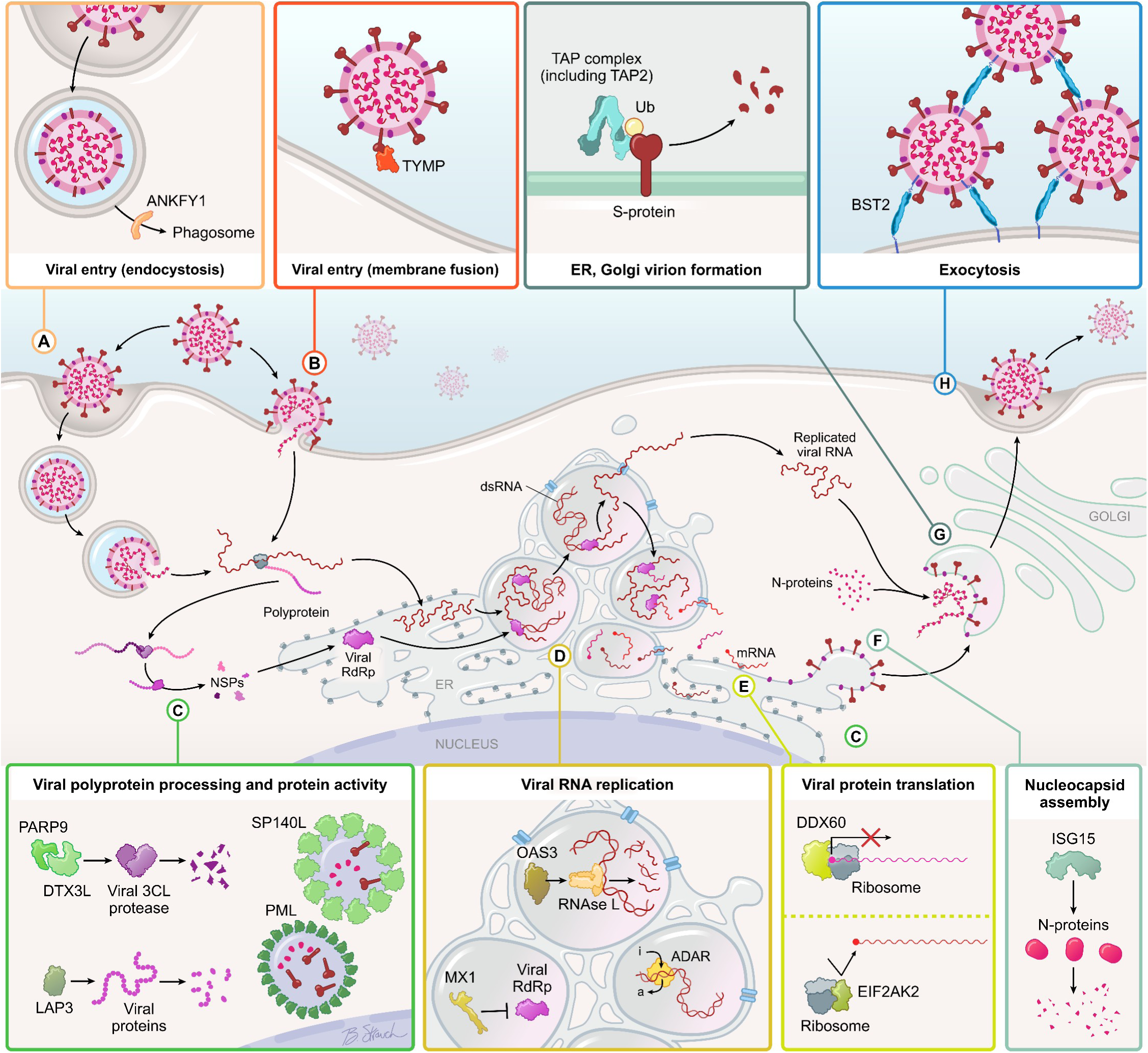
Putative Human Protein Actions Opposing the SARS-CoV-2 Life Cycle. The viral life cycle is depicted with highly expressed proteins that are associated with reductions in Neurocognitive and General Sequelae counts. **(A) Endosomal Entry.** ANKFY1 protein is associated with increased trafficking of endosomes containing other types of viruses. In the case of SARS-CoV-2, elevated ANKFY1 protein is associated with decreased post-viral sequelae counts. No mechanism specific to SARS-CoV-2 is published, but ANKFY1-mediated endosomal transport of SARS-CoV-2 may lead to phagosomal fusion and viral particle destruction similar to PML and SP140L nuclear bodies that sequester viral proteins and peptides. **(B) Membrane Entry** The major route of viral entry for SARS-CoV-2 involves membrane binding via spike protein. TYMP protein may be secreted and bind spike protein impeding membrane entry. **(C) Viral Polyprotein Processing and Protein Activity.** DTX3L and PARP9 proteins form complexes that degrade coronavirus 3C proteases which process viral polyprotein to release non-structural proteins that enable viral replication. LAP3 protein is an aminopeptidase known to restrict HIV-1. Activity against viral proteins in SARS-CoV-2 seems a likely potential mechanism. (**D) Human Opposition of Viral RNA.** OAS3 and OAS2 proteins activate human RNase L to degrade RNA with multiple viruses. With SARS-CoV-2, MX1 may inhibit viral RNA dependent RNA polymerase (RdRp). ADAR protein edits dsRNA, removing an amine group from adenosine and converting it to inosine. ADAR editing prevents auto-immunity and impedes viral RNA replication and translation. **(E) Human Ribosome.** Controlling the human ribosome controls viral translation and prevents viral takeover of intracellular machinery. EIF2AK2 protein complexes with ribosomal proteins to block 5’ entry of viral RNA. DDX60 protein complexes block 3’ exit of viral RNA, possibly freezing ribosomes in unusable states. **(F) Viral Nucleocapsid Protein.** ISG15 labels nucleocapsid proteins for degradation, blocking viral assembly. **(G) Protein Trafficking.** Proteins involved in trafficking human proteins from endoplasmic reticulum to Golgi apparatus may form complexes that bind, ubiquitinate or otherwise label viral proteins and divert them to degradation or facilitate MHC-1 presentation of viral peptides to cytotoxic T-cells. **(H) Viral Exocytosis.** BST2 protein, forms a hydrophilic loop anchored on two hydrophobic ends embedded in the human cell membrane. During exocytosis, incorporation of one BST2 protein anchor leaves a new viral particle tethered to the human cell. Multiple layers of tethered viral particles have been observed. The strategy prevents viral dissemination. Published with permission: © 2026 byte-sci creative LLC; Artist: B. Strauch.

**Table 2.** Putative Actions of Human Proteins Associated With Protective Effects Against Future Post Viral Sequelae.

| **Viral Life Stage** | **Protein (Uniprot)** | **Actions Consistent with Benefits** | **Outcomes Affected** |
| --- | --- | --- | --- |
| No Specific Stage | LGAL3BP (Q08380)<sup>41</sup> | Upregulates type 1 IFN production. | NC |
|  | STAT2 (P52630)<sup>38-40</sup> | Upregulates IFN, ISG production | GS |
| Viral Membrane Fusion Entry | TYMP (P19971)<sup>42,43</sup> | Binds viral spike protein inhibiting membrane fusion entry | NC |
| Viral Entry by Endocytosis | ANKFY1 (Q9P2R3)<sup>44,45</sup> | May modify virus endosome fusion by redirecting virus containing endosomes to fusion with phagosomes | GS |
| Viral RNA Replication | ADAR (P52948)<sup>17,68-70</sup> | A to I dsRNA Editing | NC, GS |
|  | IFIT1 (P09914)<sup>92</sup> | Inhibits NSP16, viral RNA capping | NC, GS |
|  | IFIT5 (Q13325)<sup>93</sup> | Binds viral RNA, reduces replication | GS |
|  | IRF9 (Q00978)<sup>17,38,94</sup> | Complex with ADAR to edit RNA; amplifies IFN induced ISGs | NC |
|  | MX1 (P20591)<sup>60</sup> | Inhibits viral RDRP | NC, GS |
|  | OAS2 (P29728)<sup>62</sup> | Activates RNase L degradation of dsRNA | NC, GS |
|  | OAS3 (Q9Y6K5)<sup>62</sup> | Activates RNase L degradation of dsRNA | NC |
|  | RIGI (O95786)<sup>36,37</sup> | Detects cytosolic viral dsRNA. Initiates IFN, ISG cascades | GS |
|  | STAT1 (P42224)<sup>17,38-40</sup> | Complex with ADAR edits RNA; JAK-STAT cascades. | NC, GS |
| Viral Protein Translation | DDX60 (Q8IY21)<sup>63,64</sup> | Blocks 3' viral RNA translation | NC, GS |
|  | EIF2AK2 (P19525)<sup>37,65-67</sup> | Blocks 5' viral RNA translation | NC, GS |
| Viral Polyprotein Processing and Protein Activity | DTX3L (Q8TDB6)<sup>47,48</sup> | Complexes with PARP9 to degrade viral 3C proteases | NC |
|  | HLA-B (P01889)<sup>56</sup> | Presents spike protein-derived antigens to CD8 positive T-cells | GS |
|  | LAP3 (P28838)<sup>49</sup> | Cytosol aminopeptidase may degrade viral proteins. | NC, GS |
|  | PARP9 (Q8IXQ6)<sup>47,48</sup> | Complex with DTX3L to degrade viral 3C proteases | NC |
|  | PML (P29590)<sup>51</sup> | Sequesters viral proteins in PML-nuclear bodies | NC |
|  | RNF213 (Q63HN8)<sup>53,54</sup> | Ubiquitinates viral proteins promoting degradation. | NC |
|  | SP140L (Q9H930)<sup>52</sup> | Sequesters viral proteins in SP140-nuclear bodies | NC, GS |
|  | TAP2 (Q03519)<sup>59</sup> | May help transport viral antigen for ubiquitination and degradation | NC |
|  | TAPBP (O15533)<sup>57,58</sup> | Facilitates viral peptide-MHC-I presentation to cytotoxic T-cells | NC |
|  | TRIM22 (Q8IYM9)<sup>50</sup> | Involved in degradation of NSP8, a component of viral RDRP | NC, GS |
| Nucleocapsid Assembly | ISG15 (P05161)<sup>50</sup> | ISGylates nucleocapsid protein inhibiting viral assembly | NC, GS |
|  | MX1 (P20591)<sup>18</sup> | Sequesters nucleocapsid proteins in perinuclear complexes | NC, GS |
|  | TRIM21 (P19474)<sup>55</sup> | Ubiquitinates Nucleocapsid Protein for host proteasome degradation | NC, GS |
|  | TYMP (P19971)<sup>42,43</sup> | Binds viral spike protein inhibiting final viral assembly | NC |
| Viral Exocytosis | BST2 (Q10589)<sup>46</sup> | Tethers new viral particles to the cell membrane preventing propagation | NC, GS |
| Viral Evasion | IFI35 (P80217)<sup>95</sup> | May prevent RIG-I suppression by SARS-CoV-2. | NC |

**Abbreviations:** ADP adenosine diphosphate; AMP adenosine monophosphate; dsRNA double stranded RNA; ER Endoplasmic Reticulum; GS General Sequelae counts; IMP inosine monophosphate; MHC major histocompatibility complex; NC Neurocognitive Sequelae counts; NSP non-structural protein; RDRP RNA dependent RNA polymerase; RNA ribonucleic acid.

## Discussion

We tested the hypothesis that acute respiratory viral infections initiate post-viral syndromes. We used saliva samples from volunteers seeking emergency care during the SARS-CoV-2 pandemic. By analyzing quantitative salivary proteomics, we found evidence that protein signatures expressed during acute infection are strongly linked to counts of General Sequelae that are common in Post-Acute Infection Syndrome ( **PAIS**) and to counts of Neurocognitive Sequelae that are specific to Post-Acute Sequelae of COVID19 (**PASC**).

We found that hundreds of proteins derived from Interferon Stimulated Genes (**ISG**s) exhibited clinical responses to acute infection, but stringent adjustments to reduce the rate of statistically significant but false discoveries reduced the number to 44 proteins. Among these 44 proteins, we found 29 that participated in signatures predictive of PAIS and PASC. High expression of 13 of the 29 proteins predicted low numbers of both General Sequelae and Neurocognitive Sequelae, suggesting a shared core response to viral infection. High expression of the other 16 proteins predicted either low General or low Neurocognitive Sequelae, suggesting that human defensive responses elicit different signatures depending on the specific viral infection. Based on literature review, the 29 proteins fell roughly into two groups: **(1)** those that modify human anti-viral responses by initiating or amplifying interferon responses, and **(2)** those that directly oppose one or more viral life cycle steps (**Table 2** and **Figure 3**).

Four proteins modify human anti-viral responses. RIGI detects cytosolic double stranded RNA (**dsRNA**) for many viral pathogens and activates multiple protein cascades that amplify production of interferons and other anti-viral responses.^36,37^ STAT1, STAT2, LGAL3SBP each enhance IFN production.^38–41^ All four proteins increase expression of other ISG derived proteins including the 44 clinically responsive proteins and some others among the hundreds that had weaker but suggestive responses. Whether these four, perhaps with other antiviral response modifiers, elicit specific defensive signatures matched to other viral infections such as Influenza poses a compelling question for further investigation.

Several proteins appear to oppose viral entry and exit from human cells. If secreted into the extracellular space, human TYMP^42,43^ could bind SARS-CoV-2 spike protein and hinder membrane fusion entry of virus. If present within the Golgi apparatus, TYMP could bind spike protein and block the final assembly of newly manufactured viral particles. ANKFY1^44,45^ may impede endosome mediated viral entry by redirecting virus-containing endosomes toward phagosomes. BST2^46^ hinders viral exocytosis by tethering new viral particles to the cell membrane, preventing escape and presumably awaiting cytotoxic T-cells to clear both the infected cell and newly assembled but tethered viruses.

At least 10 of the 29 human proteins inhibit viral proteins using several strategies. These include proteins that degrade (DTX3L,^47,48^ LAP3,^49^ PARP9,^47,48^ TRIM22^50^), sequester (PML,^51^ SP140L^52^) or label viral proteins for degradation after ubiquitination (RNF213,^53,54^ TRIM21,^55^ TRIM22^50^) or ISGylation (ISG15^50^). HLA-B,^56^ TAPBP,^57,58^ and TAP2^59^ cooperate in human cell surface complexes with other proteins to present peptides resulting from viral protein degradation to cytotoxic T-cells.

At least 11 of the 29 identified human proteins inactivate viral RNA using multiple strategies. Among several antiviral activities, MX1 inhibits RNA dependent RNA polymerase,^60,61^ a viral protein responsible for creating viral dsRNA, an intermediate form necessary for reproducing the single stranded RNA contained in infectious viral particles such as SARS-CoV-2. Protein variants OAS2 and OAS3 bind viral dsRNA and synthesize 2’-5’ oligoadenylates to activate human RNase L which degrades RNA.^62^ DDX60^63,64^ and EIF2AK2^37,65–67^ bind ribosomes and restrict viral RNA translation to stop production of viral proteins important for replication such as RNA dependent RNA polymerase.

ADAR stood out in our results, because it had the strongest protective incidence ratio against Neurocognitive Sequelae for patients with SARS-CoV-2 infection (**Figure 2D**) and the greatest beneficial incidence ratio against General Sequelae for all patients (**Figure 2E**). ADAR plays a central role in regulating innate immune responses by removing an amine group from adenosine incorporated in human cellular dsRNA.^68^ Deamination converts adenosine to inosine which introduces structural disruptions that impede recognition by dsRNA immune sensors, preventing aberrant immune activation and autoimmune disease.^69^ ADAR exists as two isoforms with distinct expression patterns: p150, the full-length protein, is induced by interferon signaling, while a shorter isoform, p110, is constitutively (or continuously) expressed.^70^ In different RNA viral infections, ADAR has variable effects depending on the balance between immune suppression to prevent autoimmunity and hyper - editing to deactivate viral RNA.^17^

The protective role of ADAR in our study suggests a potential contribution of immunogenic dsRNA (of viral or endogenous cellular origin) to post-acute sequelae. Low ADAR expression may allow viral RNA replication that directly leads to more severe acute disease and post-infection impairments. Alternatively, low ADAR activity may inadequately prevent human autoimmune effects^69^ secondarily triggered by SARS-CoV-2, other viral infections or the human anti-viral response itself. The latter alternative may help explain how some patients with mild initial viral infections still develop severe PAIS or PASC.

These alternatives may not be exclusive and may involve other anti-viral proteins in more intricate ways. Sufficient anti-viral signature protein activities (including those by ADAR) and ADAR-specific auto-immune controlling activities may combine to completely resolve viral infection (**Figure 4**). However, anti-viral responses that fail to completely clear viruses might combine in variable proportions with inadequate ADAR expression that allows smoldering autoimmunity to produce post-acute sequelae with a wide range of severity and divergent clinical characteristics. Ineffective viral clearance and persistent infection-triggered autoimmunity compete as possible alternative underlying mechanisms for established PAIS and PASC,^2,71^ but our results suggest that further investigations might consider interweaving the two hypotheses.^3,6^

**Figure 4.**
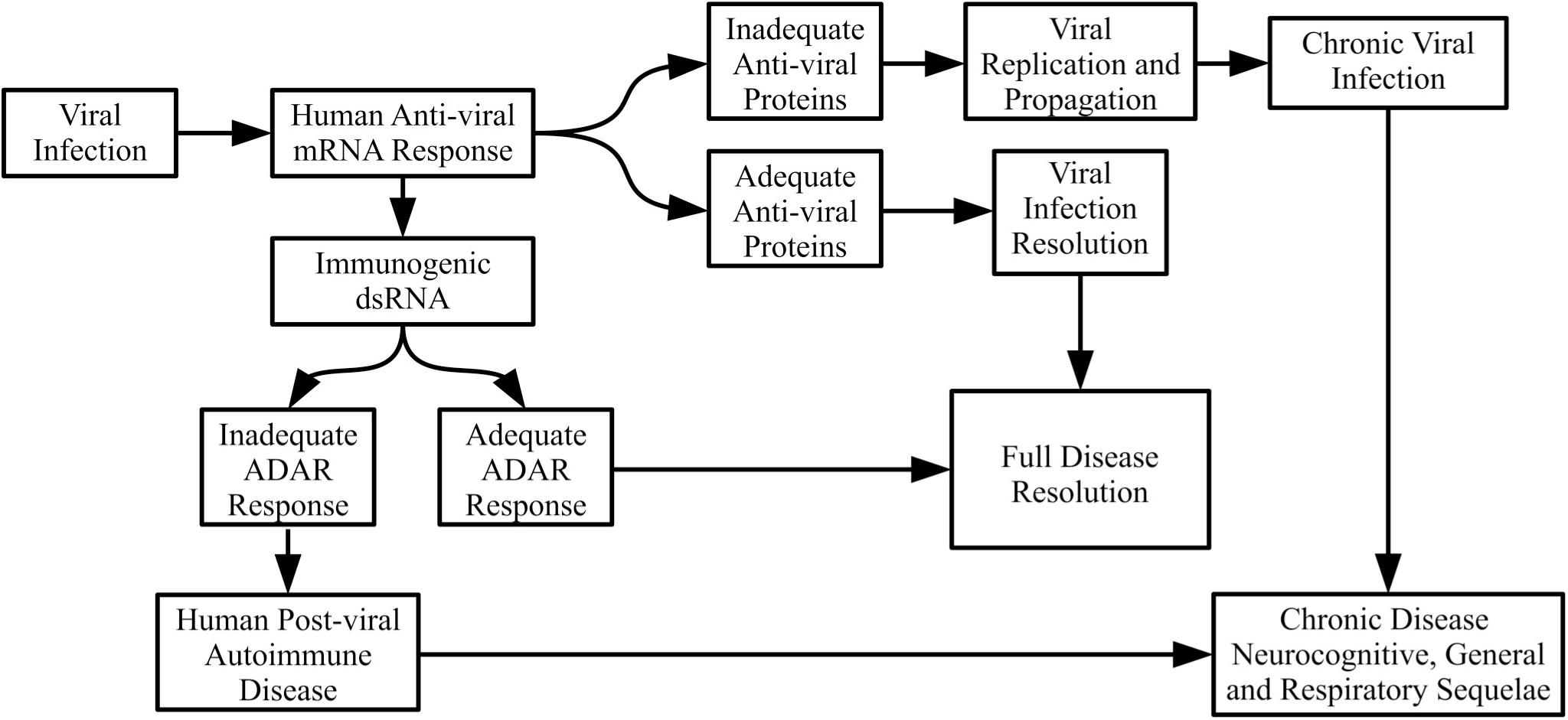
Potential Pathways from Viral Infection to Chronic Post-Viral Disease. Viral infection triggers ISG gene transcriptions leading to production of Human Anti-viral mRNA. Along the top path, Inadequate Anti-viral Proteins may lead to Chronic Viral Infection and ultimately to Chronic Disease. Adequate Anti-viral Proteins lead to Viral Infection Resolution. To the left, the Human Anti-viral mRNA Response may lead to increased Immunogenic dsRNA. If there is an Inadequate ADAR Response, Human Post-viral Autoimmune Disease may develop, contributing to post-infection Chronic Disease. Adequate ADAR Response prevents autoimmune disease and combined with Viral Infection Resolution leads to Full Disease Resolution. Combining different degrees of ADAR Response and Anti-viral Proteins may lead to varying degrees of Chronic Disease.

### Limitations of the Study

We prospectively enrolled participants with extensive past medical conditions (**Table 1**) which threatened to create disease specific bias, because patients with many past medical conditions had the lowest SARS-

CoV-2 infection rates, while patients without prior medical conditions had the highest (**Figure 2A**).^30^ In the first phase of analysis, we identified clinically relevant proteins without considering SARS-CoV-2 infection (**Figure 1A-C**) which avoids the disease specific bias. In the second phase of our analysis, uneven distribution of comorbidities relative to viral infection status threatened to distort relationships between acute-phase salivary protein expressions and post-viral sequelae, had we ignored past medical histories. We addressed potential disease specific bias by including extensive clinical annotations to mitigate confounding^72^ in our models of post-acute sequelae. These procedures enabled successful identification of antiviral proteins that were beneficial against post-acute sequelae (**Figure 2D and 2E**).

The lack of significant associations between acutely expressed proteins and counts of Respiratory Sequelae was surprising, because these were commonly reported sequelae (**Table S1**). Linear regression models showed similarities in protein-associated incidence ratios derived from Respiratory Sequelae and General Sequelae count models, but the respiratory sequelae and individual protein models lacked statistical power (**Figure 2H**). Understanding protein relationships with Respiratory Sequelae may simply require a larger study cohort.

Our study involved a single center, potentially limiting generalizability, however, we had few refusals to participate, and we had substantial representation by every racial and ethnic group commonly tracked in the US. The types and densities of living conditions among our volunteers suggest that we included individuals from a range of socioeconomic groups. Our models yielded conclusions about the pathobiology of post-acute sequelae that remained stable in sensitivity models testing the effects of race, ethnicity and socioeconomic status.

We did not ascertain SARS-CoV-2 variants to understand strain-specific potential impacts. However, our 28-month recruitment period began during SARS-CoV-2 α-variant infection waves, spanned all δ-variant waves and continued through several waves of ο-variants.^33^ Our sensitivity analyses demonstrated stable models regardless of enrollment date, implying that our results depend on the constancy of human anti-viral defenses rather than continuously evolving SARS-CoV-2 genomes.

## Methods

### Clinical Data Acquisition

#### Patients

With University of Utah Investigational Review Board and Biosafety Committee approvals (IRB_134424), we recruited a cohort from the University of Utah Emergency Department (ED) who received PCR-based testing for symptoms and signs consistent with a viral syndrome between 11 January 2021 and 9 May 2023. We included adults for whom ED staff ordered PCR testing for SARS-CoV-2, who provided written informed consent in English, and who agreed to provide a saliva sample. We recruited patients beginning mid-morning and ending mid-day, ∼4-5 hours, Monday to Friday to match the highest daily volume of patients and to limit times between collection and sample processing to less than 4 hours.

We excluded patients unable to produce saliva, were without viral syndrome complaints or findings, received routine precautionary testing for non-infection admission reasons, were prisoners or were unable or unwilling to provide consent. Because we tested the hypothesis that acute infections initiate post-viral syndromes, we excluded patients from analysis who had symptoms beginning more than 30 days before enrollment. Using a standardized questionnaire, we recorded the nature and duration of acute symptoms and the dates of prior SARS-CoV-2 vaccinations, infections and treatments, open-label or blinded study participation, acute evaluation findings and same-day outcomes such as hypoxemia, ED discharge or hospital admission.

We reviewed charts at the time of acute illness and periodically through 18 December 2025. We manually opened individual charts, read all human-generated notes and searched for indications of neurocognitive dysfunction (brain fog, confusion, inability to make decisions, depression, anxiety, specific nerve impairments such as loss of taste or smell, headaches, seizures, transient ischemic attacks, cerebrovascular accidents), general long term symptoms (fatigue, post-exertional malaise, disturbed or altered sleep, orthostatic intolerance, nausea, vomiting, diarrhea, poor appetite, chills, hot and cold spells, joint or muscle pains, palpitations, renal disease, changes in abilities to live life including discontinuation or reductions in work or school, difficulty with or new or increased needs for assistance with activities of daily living), respiratory issues (persistent need for newly prescribed or persistently increased oxygen supplementation, dyspnea, exercise-associated dyspnea, chest pain, cough, increased cough) or any other impairments consistent with PASC or PAIS.^3,8–10,15^ We noted whether any findings started before, with or after the enrollment encounter.

We counted ED, clinic and hospitalization visits as indirect quantitative indicators of healthcare needs for the years preceding and following enrollment for acute illness. Finally, we noted deaths from any cause. We censored follow-up of all patients at the time of last contact, death or the end of follow-up on 18 December 2025. We collected and managed all clinical data using the Research Electronic Data Capture (REDCap) tools hosted at the University of Utah.^73,74^

#### Saliva Samples

From each patient, we requested a 5 ml saliva sample and allowed up to 20 minutes for collection in a sterile 50 ml conical tube (ThermoFisher Scientific, Waltham, MA, USA) which we labeled and placed on ice for transport for laboratory processing within 4 hours of collection. Saliva samples were mixed 1:1 with Hanks buffered saline solution (ThermoFisher) and vortex mixed after 1:1 dilution with RNALater (ThermoFisher). After division into approximately 0.5 ml aliquots, samples were centrifuged for 25 minutes in 1.5 ml vials (ThermoFisher) in a Beckman Coulter Microfuge 18 at 14,000 RPM (18,000× *g*) (Beckman Coulter Life Sciences, Indianapolis, IN, USA). Supernatants were transferred to fresh vials and stored at -80 °C until use. Frozen supernatant aliquots were used for multiplex ELISA assays. Pellets were flash frozen using a dry ice-methanol bath and stored at -80 °C until use. Pellet aliquots were used for protein and RNA preparations. All aliquots were used following a single thaw.

### Laboratory Data Acquisition

#### Protein Preparation for Mass Spectrometry

Thawed saliva pellet fractions were lysed in RIPA buffer, and protein was precipitated with 9× volume of ice-cold acetone and resuspended in 1× S-TRAP buffer (50 mM Triethylammonium bicarbonate [TEAB, Sigma Aldrich, St Louis, MO, USA], pH 8.5 and 5% sodium dodecyl sulfate [Sigma Aldrich]). Total protein content was quantified using the Pierce™ BCA Protein Assay Kit (ThermoFisher) according to the manufacturer’s instructions; 10-15 µg of protein from each sample was diluted into 25 µL of 1× S-TRAP buffer for submission to the University of Utah Proteomics Core Laboratory Facility.

In the Proteomics Core Laboratory, proteins were reduced with 20 mM dithiothreitol at 37 °C for 30 minutes. Samples were alkylated with 40 mM iodoacetamide (Sigma Aldrich) at room temperature for 45 minutes in the dark and acidified with 55% phosphoric acid (Sigma Aldrich) to a final concentration of 4.5% phosphoric acid. We added 330 µl of binding buffer (10% 100 mM TEAB [Sigma Aldrich], final pH 7.5 with phosphoric acid [Sigma Aldrich], and 90% MeOH [Honeywell Research Chemicals, Morris Plains, NJ, USA]) to each sample. We vortex-mixed samples and added them directly to S-TRAP columns (Protifi, Fairport, NY, USA). The columns were centrifuged at 4000× g for 1 min at room temperature. Each column was washed 3× with 150 µl of binding buffer, and each washed column was transferred to a new clean tube. Protein was digested with 0.75 µg of trypsin/Lys C (Promega Corporation, Madison, WI, USA) for 2.5 hours at 47 °C. The peptides were eluted from each column with 40 µl of 50 mM TEAB pH 8.5 followed by 40 µl of 0.2% formic acid (ThermoFisher) and 40 µl of elution buffer (50 mM TEAB pH 8.5 + 0.2% formic acid + 50 % acetonitrile [ThermoFisher]). The peptides were dried to completion and resuspended in 300 µl of 0.1% trifluoroacetic acid (Sigma Aldrich) and desalted using Pierce™ Peptide Desalting Spin Columns (ThermoFisher) according to the manufacturer’s instructions. The peptides were resuspended in 35 µl of 0.1% formic acid, and total peptide amount was determined using Pierce™ Colorimetric Peptide Assay Kits (ThermoFisher). All samples were normalized to 0.02 µg/µl in 0.1% formic acid for liquid chromatography-(LC)-MS/MS analysis.

#### Mass Spectrometry

Reversed-phase nano-LC-MS/MS was performed on a nanoElute 2 (Bruker Daltonics, Billerica, MA, USA) coupled to a timsTOF-MS Pro2 mass spectrometer equipped with a nanoelectrospray source (Bruker Daltonics). We injected 120 ng of each sample directly onto the LC reverse-phase ReproSil C18 150 mm × 0.15 mm nanocolumn (Bruker Daltonics) heated to 50 °C. The peptides were eluted with a gradient of reversed-phase buffers (Buffer A: 0.1% formic acid in 100% water; Buffer B: 0.1% formic acid in 100% acetonitrile [Sigma Aldrich]) at a flow rate of 0.5 µl/min. The LC run lasted for 45 minutes with a starting concentration of 5% buffer B increasing to 28% buffer B over 38 minutes, up to 35% buffer B over 2 minutes and held at 95% B for 5 minutes. The separation column was equilibrated with 4 column volumes at 800 bar following each run. The mass spectrometer was operated in parallel accumulation serial fragmentation (PASEF) data-independent acquisition (DIA) MS/MS scan mode to analyze each experimental sample. The trapped ion mobility spectrometry section was operated with a 120 ms ramp time at a rate of 7.93 Hz and an ion mobility scan range of 0.6-1.4 V·s/cm^2^. DIA-PASEF window parameters were set to mass width of 25 Da and 47 mass steps/cycle. MS and MS/MS spectra were recorded from 147 to 1,322.6 m/*z*. Samples were run in alternating group order to reduce batch effects.

#### Mass Spectrometry Data Analysis

We calculated abundance based on peak intensity (spectral intensity) for each protein using the DIA-NN version 1.8.1 software^75^ and the human peptide spectral library (Bruker Daltonics) uploaded with 513,182 peptides. We allowed for 1 missed cleavage following trypsin/Lys C digestion but no fixed modifications. The variable modifications of methionine oxidation, N-terminal acetylation and cysteine carbamidomethylation were considered with a mass tolerance of 15 ppm for precursor ions and a mass tolerance of 10 ppm for fragment ions. We filtered results with an FDR^25^ of 0.05 for both proteins and peptides. A minimum of 1 unique peptide was reported for each protein identified.

#### Multiplex ELISA Assays

Using LEGENDplex™ Hu Anti-Virus Response Panel 1 13-plex ELISA kits (Biolegend, San Diego, CA, USA), we assayed interferon-(IFN)-λ_1_ (interleukin-[IL]-29), IL-1β, IL-6, tumor necrosis factor-(TNF)-α, IP-10, IL-8, IL-12p70, IFN-α_2_, IFN-λ_2_ (IL-28A), granulocyte macrophage-colony stimulating factor (GM-CSF), IFN-β, IL-10 and IFN-γ in the supernatant fractions. We modified manufacturer instructions by adding 6 μl 0.0825% electron microscopy grade glutaraldehyde to all experimental and standard curve wells (final concentration 0.00825%, catalog number 16216, Electron Microscopy Science, Hatfield, PA, USA) and incubated at 4 °C for 2 hours to reduce the risk of infection of laboratory personnel. Two hours is 4 times longer than needed to reduce the SARS-CoV-2 50% cell culture infectious dose from 10^6.5^ plaque forming units (PFU)/ml to below limits of detection (∼6 PFU/ml) verified in triplicate using Vero E6 and Calu-3 cell cultures, as determined at The Institute for Antiviral Research, Utah State University (unpublished data). We assayed the 96 well plates using a Canto flow cytometer which limits sample aerosolization^76^ (BD Biosciences, San Jose, CA, USA) in the University of Utah Flow Cytometry Core Laboratory Facility.

#### RNA-sequencing

We purified RNA from saliva pellets using *Quick*-RNA Miniprep Kits according to manufacturer instructions (Catalog R1055, Zymo Research, Irvine, CA, USA). We used the variation for RNA purification at indicated points in the instructions. RNA concentrations were measured using A260/A280 ratios in a NanoDrop Lite microvolume spectrophotometer (ThermoFisher). We diluted samples as needed with molecular grade water from a MilliQ purification system (Millipore, Sigma, St Louis, MO, USA) to achieve the concentration needed for submission to the University of Utah DNA Sequencing Core Laboratory Facility.

To the DNA Sequencing Core Laboratory, we submitted 100 ng of purified RNA per sample for sequencing in 18 uL of molecular grade water (MilliQ purification system, Millipore). We processed the RNA by following the Watchmaker Genomics RNA Library Prep with Polaris Depletion protocol (kit number 7BK0002-096, Watchmaker Genomics, Boulder, CO, USA) which includes steps using 80% ethanol made fresh daily in the Core Laboratory using molecular grade water and 100% ethanol (Decon Labs, Inc, King of Prussia, PA, USA). The libraries were built per Library Construction Protocol A (Watchmaker Genomics) with 1 minute RNA fragmentation at step A4.5. Full length IDT X-Gen UDI-UMI adapters (part number 10005903, Integrated DNA Technologies, Inc, Coralville, Iowa, USA) were prepared with low EDTA TE, pH 8.0 (cat. 351-324-721, Quality Biological, Gaithersburg, MD, USA) and used with 1 μM adapter solution (Integrated DNA Technologies) when adding the adapters/barcodes. Post construction, libraries were subjected to 12 cycles of PCR amplification using an Applied Biosystems 4483636 ProFlex Base Thermal Cycler (ThermoFisher). All samples were converted to run on the Element Biosciences AVITI24 sequencer (Element Biosciences, San Diego, CA, USA) per the Element Adept Library Compatibility Workflow Adept Rapid PCR-Plus Protocol using 5 cycles. The libraries were checked for size using a 12 capillary AATI Fragment Analyzer (discontinued model, Agilent, Santa Clara, CA, USA) using the High Sensitivity NGS kit (part number DNF-474-0500, Agilent) and quantitated using the Qubit High Sensitivity kit (part number Q33231, ThermoFisher). Libraries were diluted to 1 nM with molecular grade water, and equal aliquots were taken of each library in sets of 20 per High Output 2×150 Cloudbreak (Element Biosciences) flow cell.

We used Kraken2 scripts to classify taxonomic sequence outputs.^28^ We focused on human respiratory viral pathogens including *Betacoronavirus pandemicum* (SARS-CoV-2) to augment PCR identification of individuals with infections by SARS-CoV-2 at the time of sample collections. Using samples from patients who had PCR testing the same day as saliva collection, we evaluated the relative sensitivity and specificity of PCR and RNA-Seq. We classified patients as SARS-CoV-2 positive for our study by either PCR or RNA-Seq detection because of the low likelihood of false positive combined testing.

### Statistical Analyses

#### Software

We used the R statistical environment for all data processing and analysis^77^ with additional packages including chron, data.table, future, future.apply, MASS, mgcv, rlist and stats.^32,78–83^ We generated all of the R code used in this project without the assistance of artificial intelligence.

#### Patient Characteristics

We performed descriptive analyses of clinical characteristics and acute and longterm outcomes. Because our patients were recruited by convenience, we specifically evaluated whether pre-enrollment medical conditions were associated with the likelihood of SARS-CoV-2 infection. We calculated principal components^29^ (**PC**) based on all past medical history variables and used the first PC (**MedPC1**) as the independent variable in logistic regression^31^ with SARS-CoV-2 detection as the dependent variable to test for the presence of bias.

#### Protein Measurements by timsTOF-MS

Protein non-detection by operating a mass spectrometer in PASEF DIA mode indicates measurement below the level of detection which provides partial information. We excluded proteins from further analyses that were fully informative for fewer than 30 patients. We log-transformed MS spectral readouts because they were positive and approximately symmetrically distributed after transformation. We used base 2 logarithms to enhance the ability to interpret statistical models using these values. To use partial information, we examined fully informative log_2_ transformed data using quantile plots^27,84^ of the data against a normal distribution.^84^ We fitted a generalized additive model^83^ to determine minimum detected values which we defined as the lowest sample value within 3 standard deviations of its fitted value. For any spectral intensities that were below the identified minimum or were only partially informative, we assigned the value of the minimum detected level × 0.99 to minimize potential bias from arbitrary value assignments.^85,86^

#### Protein Measurements by Multiplex ELISA

We derived raw multiplex ELISA protein measurements from standard curves using Biolegend software (Biolegend) which we log_2_-transformed before analysis. Multiplex analysis conserves sample and reduces assay costs, however, it requires a single dilution that is optimized on average but which may increase the number of values with only partial information with some values determined to be below and others to be above detection limits for specific analytes. Similar to partially informative timsTOF-MS measurements, we assigned the value of the minimum detected level × 0.99 or the maximum detected level × 1.01, for partially informative values below the minimum or above the maximum detected values, respectively, to reduce potential bias from arbitrary value assignments.^85,86^

#### Data Exploration and Quadratic Regression

Due to the large number of candidate proteins for consideration, some preliminary reduction was needed prior to more formal regression analyses seeking associations between acute protein measurements in participants with and without SARS-CoV-2 infections and subsequent sequelae counts. Our timsTOF-MS and multiplex ELISA measurements included multiple proteins known for anti-viral activity.^18–22^ We previously studied several that were upregulated in the setting of SARS-CoV-2 infection suggesting the possibility of a fold-change analysis to choose proteins for further study.^87^

However, we intended to investigate the role of SARS-CoV-2 in the development of PASC, thus we could not also use detection of SARS-CoV-2 and fold-change protein responses for reducing candidate protein numbers. Such a selection procedure invalidates the statistical properties underpinning the formal regression analysis. More generally, serious issues of statistical calibration arise when essentially the same scientific question is posed in different guises, the answer to the first forming the basis for subsequent analyses.^23^

The data collected by this study included a large amount of clinical information including time since first symptom which was not available in our previous study.^87^ These data allowed exploration^27^ of protein behaviors during the first 30 days after first symptom. To explore, we reviewed plots of every protein measured by tims-TOF-MS or multiplex ELISA as the dependent variables against days since first symptom, which is associated with estimated time of infection, as the independent variable. Observing first that multiple anti-viral proteins including those that we studied previously^87^ followed a non-linear rising and falling pattern suggestive of a response to acute illness, we performed quadratic regression analysis^24^ for every measured protein with the model,

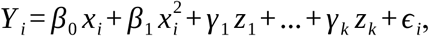

where for each participant *i, y_i_*is the log_2_-transformed protein measurement, *β_0_* and *β_1_* are the linear and quadratic term regression coefficients, respectively for *x_i_*, the number of days since first symptom, *γ*_1_..*γ_k_* are coefficients for other covariables *z*_1_..*z_k_* and *ɛ_i_* is the mean-zero error term. We adjusted quadratic regression *p*-values for non-zero coefficients of the quadratic terms using FDR^25^ analysis. This first preliminary phase of analysis discovered the reduced set of proteins for the second phase of analyses that follow.

#### Evaluation of Confounding

Confounding may arise from unevenly distributed patient characteristics, so we used principal components analysis^29^ to establish the potential for disease-specific biases.^30^ After centering and scaling the data, we performed principal components analysis of intrinsic factors, age and sex, 16 comorbidities, prior year clinic, ED and hospital visit counts and initial outcomes of the ED visit, hypoxemia and hospital admission which roughly indicated severity of acute presentation.^88^ Using the first principal component which we termed MedPC1, we fitted a logistic regression model^31^ using SARS-CoV-2 detection as the dependent variable. The strong inverse association between SARS-CoV-2 and MedPC1 demonstrated the need to adjust any models involving SARS-CoV-2 with at least some comorbidities, intrinsic factors and other patient characteristics. We proceeded to logistic regression models with SARS-CoV-2 as the dependent variable and each intrinsic and past medical condition as independent variables to demonstrate that individual variables subsumed into MedPC1 were potential confounders and therefore potential adjustments for further models exploring PASC.

To gain more precise insights on potential confounders, we assessed combinations of up to seven intrinsic and comorbidity variables without interactions in negative binomial models of post-viral Neurocognitive, General and Respiratory Sequelae counts.^32^ We selected potential confounders from among intrinsic variables, age and sex and past medical conditions that affected more than 20% of the patients as independent variables. These variables included prior SARS-CoV-2 infection, hypertension, central nervous system (**CNS**), mental health, obesity, heart, lung, gastrointestinal (**GI**) and rheumatologic diseases. We included *diabetes mellitus* because of clinical importance.

We evaluated model performance using log-likelihood ratio tests^88^ and selected among potential confounder combinations giving comparable quality of model fits to maximize parsimony and clinical interpretability. For example, in Neurocognitive Sequelae models, we adjusted for mental health and CNS diseases, because these are more directly related to the outcome than alternatives such as GI and rheumatologic diseases. For General Sequelae models, we adjusted for age, obesity, and heart disease. We adjusted for lung and heart disease, obesity, cigarette smoking and acute hypoxemia for Respiratory Sequelae models.

#### Post-Viral Outcomes

We used the protein measurements identified in the first phase of analysis by quadratic regression (quadratic term *p*_FDR_ < 0.01) as the input variables of primary interest to analyze post-viral outcomes in the second phase of analysis. We analyzed the count of distinct sequelae per patient as the dependent variable in three non-overlapping classes (neurocognitive, general and respiratory symptoms).^10,15^ The counts appeared Poisson-like in distribution. To allow for the possibility of over-dispersion relative to a Poisson model, we used a negative binomial regression for the counts.^32^ In particular, our interim and final analyses produced a conditional variance that exceeded the conditional mean suggesting a negative binomial model. The negative binomial model, implicitly averaging over an unknown gamma distribution of unit-specific nuisance parameters, achieves a greater degree of robustness to the omission of relevant baseline variables. Using negative binomial models, we evaluated each protein measurement twice, because we included the SARS-CoV-2 detection variable as either an additive or interacting independent term to investigate whether protein expression levels were independent of specific viral syndrome cause or were directly related to SARS-CoV-2 infections, respectively.^89^

#### Models of Post Viral Symptom Outcomes

We calculated principal components using centered and scaled measurement values of the proteins selected for clinical relevance by quadratic regression analysis.^29^ We explored associations of the first principal component which we termed QuadPC1 as an independent variable for post-viral neurocognitive, general and respiratory symptom outcomes in negative binomial models.^32^

We evaluated SARS-CoV-2 detection status as either an additive explanatory variable or as an interacting modifier for protein effects. We then adjusted models first with principal components of past medical history used to detect disease specific biases^30^ and with the specific adjustment variables that improved clinical insights for each post-viral outcome. We used the results to select the model framework for evaluation of post-viral outcome relationships with individual proteins, specifically whether to include SARS-CoV-2 as an additive or interaction term.

To discover protein-specific details, we proceeded with negative binomial models^32^ of counts of each class of dependent post-viral symptom outcome as dependent and individual proteins and SARS-CoV-2 status adjusted by individual confounding variables as independent variables. We initially evaluated the individual proteins identified by quadratic regression modeling in univariable models and proceeded to multivariable modeling that included potential confounders as additional independent variables.

We selected past medical and intrinsic variables with unadjusted *p*-value < 0.05 for multivariable evaluations with proteins and SARS-CoV-2. We used the confounders for each class of outcome variables selected above for parsimony and clinical relevance when log-likelihood ratio testing results were of comparable quality.

#### Sensitivity Analyses

We performed sensitivity analyses of all negative binomial models to examine the impacts of race, ethnicity, housing and sample quality. We examined sensitivity of all adjusted negative binomial models to each of the medical conditions excluded from adjusted models because they affected < 20% of participants. Some patients had a period of 1 or more days between PCR testing and saliva collection. We performed sensitivity analyses of all models for effects of the time between PCR testing and saliva collection to exclude potential immortality biases. We examined the impact of date of enrollment as SARS-CoV-2 variants change constantly.^33^ Our study enrolled patients from the peak of α-variant infections, encompassed the rise and fall of δ-variant infections and included about the first year of ο-variant infections which dominated infections through the end of the study.^33^

### Synthesis

#### Reactome Pathway Expression Analyses

To understand the biological pathway organization of proteins identified by negative binomial models of the non-overlapping classes of post-viral infection impairments, we submitted results to the Reactome database for over-representation analyses.^34^ We used Uniprot^90^ protein designations with negative binomial model coefficient estimates as the quantitative element. We retained human-curated pathways with *p*_FDR_ < 0.05 in which submitted proteins numbered at least 5 or represented more than 20% of the pathway.

#### Literature Review

To better understand the extent of the immune defensive response represented by acutely elevated proteins that proved to have significant associations with subsequent post-viral outcomes, we performed an initial review of citations regarding the anti-viral activity of each protein at the Uniprot site,^90^ if available. We proceeded to a short literature review focused on progressively more general search terms. For each protein, we started with “protein AND (SARS-CoV-2 OR COVID19)” and proceeded to “protein AND SARS,” “protein AND coronavirus,” “protein AND virus,” and finally “protein mechanism.” We preferred human over animal or *in vitro* data. We stopped when we found sufficient investigative evidence to report either a known anti-viral mechanism specific to SARS-CoV-2, specific to SARS, other coronaviruses or viruses in general or found sufficient evidence in other contexts to propose a putative mechanism for significant protein and post-viral sequelae relationships.

## Data Availability

The need to protect individual level patient health information prevents sharing of all data. Where possible, we provide raw data that are deidentified. We provide software and data that produce nearly identical figures as Figures 1 and 2 and Tables S2 and S8 but differ because of the use of 2 digit age instead of exact age to protect patient privacy. Multianalyte ELISA and timsTOF-MS derived spectral intensities and the software to make Table S3, S6 and S7 produce duplicates of the information presented in those tables. Software and timsTOF-MS derived spectral intensity data with adjustments for partial information are provided that allow viewing of quadratic fits for all proteins stepping through the proteins one at a time or one screen full at a time. For all the Figures and Tables, code may be sourced inside a command line R interface. The code for viewing quadratic fits of proteins is intended to be modified by the user to allow viewing of 1 to all the proteins found in saliva in our study with commands to be copied and pasted to a command line R interface. We do not provide data or software to recreate Tables 1, S4 and S5 because of the need to protect patient privacy. The data for these tables included explicit identifiers or implicit identifiers because of the creation of categories of patients with fewer than 5. We either omitted or combined these categories to protect privacy in the manuscript. We did not write software to create Table 2 and Figures 3, 4, S1 and S2.

Raw proteomics data are not available because implicit identifiers are embedded. We do not have explicit permission from participants to share raw RNA-Seq data. Please contact the lead author to discuss access that may be possible with IRB supervision.

## Supplemental Information

Document S1. Supplemental Figures S1-S2

Figure S1. CONSORT Diagram of Patient and Data Flow through the Analysis

Figure S2. Statistical Analysis Plan

Document S2. Supplemental Tables S1-S9

Table S1. Post-viral Sequelae

Table S2. Quadratic Regression Models of Protein Quantities Dependent on Days from First Symptom

Table S3. Quadratic Regression Results for Soluble Human Saliva Proteins

Table S4. Viral Detections

Table S5. Principal Components for Intrinsic and Comorbidity Variables

Table S6. Principal Components for 44 Proteins Clinically Relevant for Viral Syndromes

Table S7. Protein Incidence Ratios Associated with Decreased Subsequent Neurocognitive Sequelae

Table S8. Protein Incidence Ratios Associated with Decreased Subsequent General Sequelae

Table S9. Reactome Pathways

## Acknowledgments

We acknowledge the foundational contributions of Professor Sir David Cox who inspired the initial overall study and design during a meeting held in March of 2020 but who passed away^91^ prior to completion. We are grateful for reviews of the manuscript by Dr. Ramsey Hachem.

## Author Contributions

All authors participated in critical reviews and revisions of the manuscript and provided insights that collectively modified study performance, analyses, interpretations and writing. TGL led the study, participated in all aspects of study performance, secured funding, wrote the initial manuscript and is first author. TGL, JK and AU recruited patients, collected samples and administered enrollment questionnaires in the Emergency Department with advice from SCH, MGO and STY. RJA and BLB provided critical understand of ADAR and rewrote pertinent sections of the manuscript. BLB provided critical guidance for figure 2 and inspiration for figure 4 with inputs from RJA, TGL and JLJ. AMW provided guidance for Table 2. TGL, HSB, CK, RDS and FRA planned and designed the study, participating in the initial meeting with Sir David and performed the statistical analyses with input from SY on patient selection methods. LGVB added key insights pertinent to proteomics analysis and interpretation beyond those provided by JEC, FBTPL and AM. JEC led and AM performed the timsTOF-MS proteomics analysis and initial protein identifications, and with FBTPL provided specific information on selected proteins and peptide sequences including the discovery of *in vivo* HLA-H translation. SG and EBT provided experimentally derived data developed at the BSL-3 Laboratory at Utah State University that allowed safe handling of samples containing live SARS-CoV-2 and other live viral human pathogens. SCH, MGO and STY provided ED access and coordinated study activities with the many physicians, nurses and staff of the University of Utah Emergency Department. MNH, YL, ABS and EZ designed and performed the laboratory handling, processing and safety protocols. YL performed the multianalyte ELISA measurements and initial protein with key assistance from MNH and RNA extractions with key guidance from MWH. JK and AU with help from TGL, JLJ and KAP performed patient recruitments and designed and managed initial sample collection procedures. JLJ and KAP oversaw regulatory procedures, managed funds, maintained sample inventory and with YL oversaw laboratory operations. MM and DW performed bulk RNA-Seq and sequence processing including Kraken2 processing. JEM directed the Flow Cytometry Core Laboratory and oversaw plate reading and processing of raw measurements to derive final protein measurements. EAM and NH coordinated and provided samples and follow up for patients admitted to the intensive care units following ED evaluation. KAP designed, edited and maintained the REDCap data collection instruments with input from TGL, JK and AU. AU designed chart review goals with input from TGL and KAP and performed reviews and interpreted chart findings. TGL and FRA led the analysis with critical inputs from HSB, CK, RDS, SKY and simplifying modifications from BCC and AT. MGO and STY were senior authors for clinical activities, and FRA was senior author for analytical activities.

