## Supplemental Figures S1-S2 for "Strong Protein Responses to Acute Infection Control General Post-Viral Sequelae and SARS-CoV-2-specific Neurocognitive Sequelae"

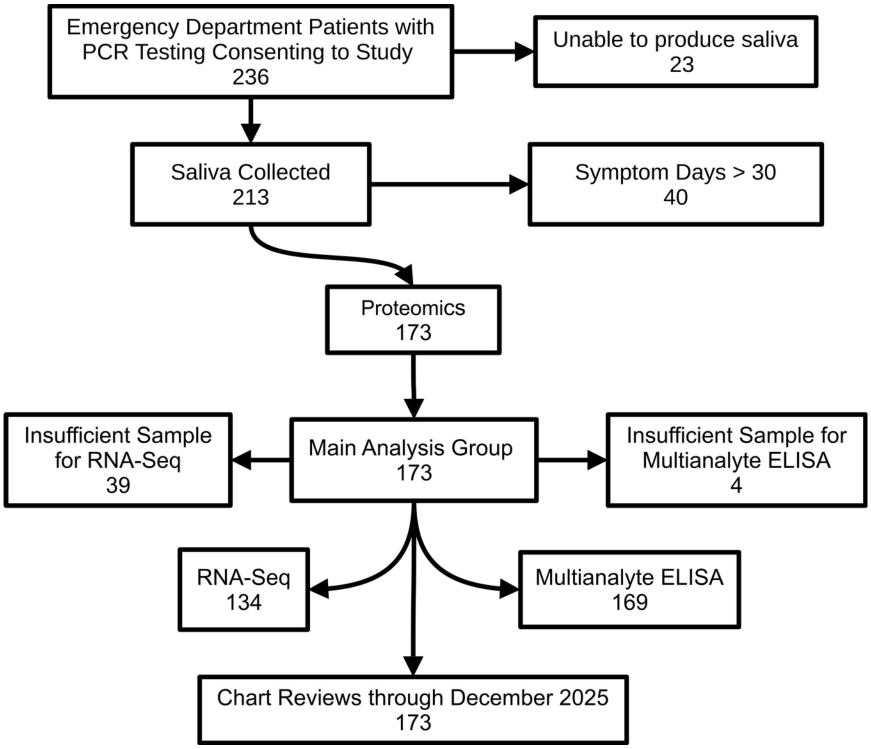


**Figure S1. Consort Diagram.** Patient and laboratory sample flow during the study. Out of 236 patients who provided written informed consent, 23 were unable to produce saliva and 40 reported that more than 30 days had elapsed since the first symptom. We performed proteomics on the 173 samples using timsTOF-MS, and we were able to perform RNA-Seq on 134 of those samples and multianalyte ELISA on 169. For all 173 participants, we performed multiple chart reviews through the end of the study in December 2025.

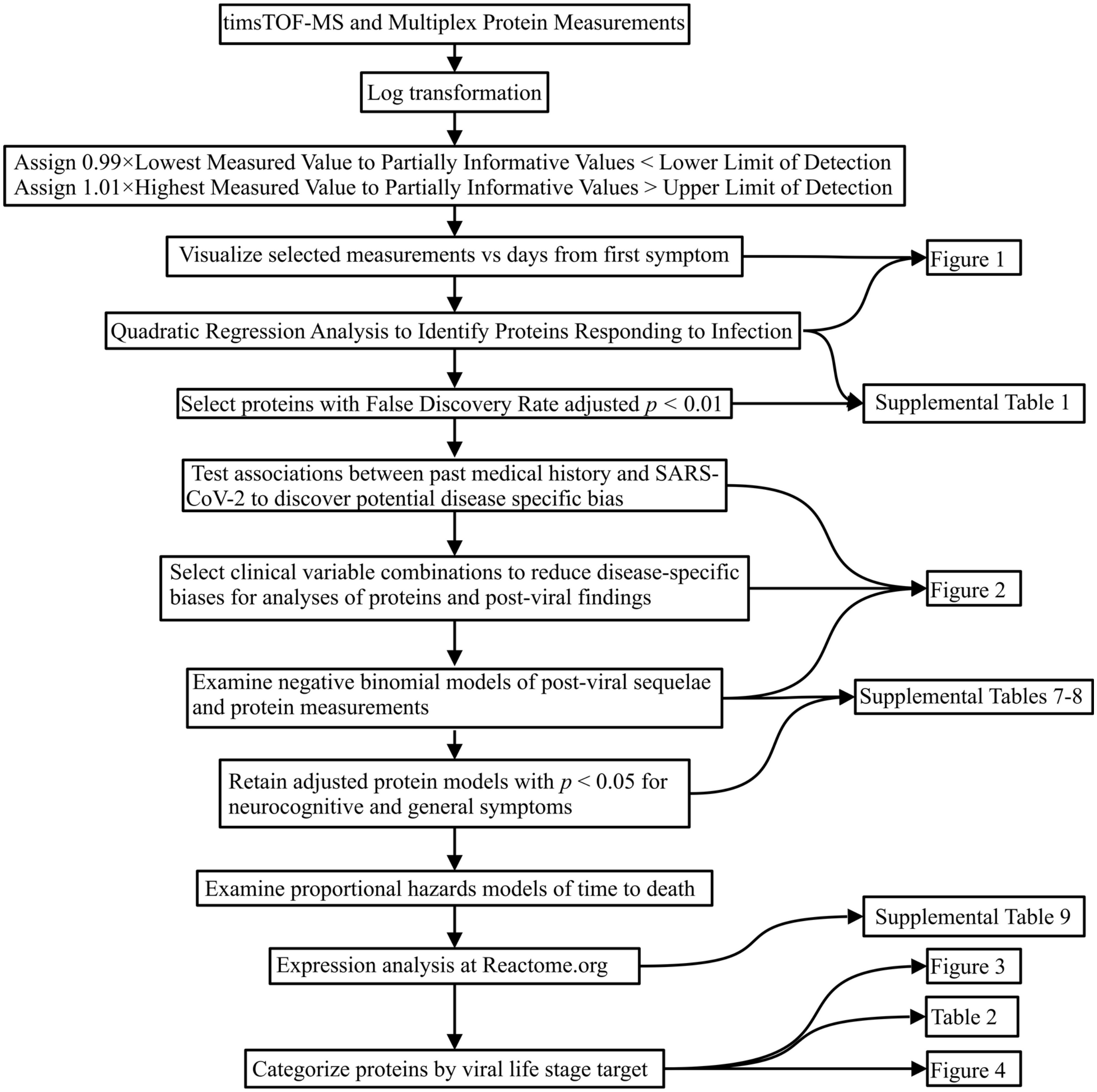


**Figure S2.** Statistical Analysis Plan. We assigned values for protein measurements that were partially informative in that values were below the range of detection for all timsTOF-MS and some ELISA assays and above the range of detection for other ELISA values. To avoid compromising statistical validity of subsequent analyses after reduction to a more manageable number of proteins, we retained proteins for further study based on a covariate-adjusted quadratic regression of each protein on the duration of disease (see model equation in **Methods** *Data Exploration and Quadratic Regression*). By focusing on clinically-relevant protein behavior for this reduction phase and using a different framing of a scientific question to that used in subsequent analysis phases, we avoided post-selection inference issues for models that assessed relationships between specific proteins and viral infections and subsequent persistent post-viral symptoms.
