## Supplemental Tables S1-S9 for "Strong Protein Responses to Acute Infection Control General Post-Viral Sequelae and SARS-CoV-2-specific Neurocognitive Sequelae"

**Table S1. Post-Viral Sequelae**[*]

| Symptom Category | Individual Sequelae | No SARS-CoV-2, n = 73 | | SARS-CoV-2, n = 100 | | Adjustment Variables for Negative Binomial Regression |
|---|---|---|---|---|---|---|
| | | Counts | Total Counts | Counts | Total Counts | |
| ***Neurocognitive*** | Brain Fog, Confusion, Taste or Smell disturbances | 6 | 33 | 16 | 45 | Mental Health and Central Nervous System Diseases |
| | Depression or Anxiety | 9 | | 10 | | |
| | Chronic Headaches and Other (paresthesias, hearing loss, double vision) | 18 | | 19 | | |
| ***Generalized*** | Chronic Fatigue | 19 | 78 | 30 | 109 | Age, Obesity, Heart Disease |
| | Sleep Disturbances | 6 | | 11 | | |
| | Persistent Nausea, Poor Appetite | 12 | | 9 | | |
| | Recurrent Joint Pains, Muscle Aches, Chills | 16 | | 27 | | |
| | Palpitations | 7 | | 11 | | |
| | Changes in Work or School | 7 | | 10 | | |
| | Other (orthostatic blood pressure changes, dizziness or falls, swelling of legs, hands or face, hair loss, skin changes) | 11 | | 11 | | |
| ***Respiratory*** | Increased Oxygen | 7 | 55 | 10 | 69 | Lung and Heart Disease, Obesity, Cigarette Smoking, Hypoxemia in ED |
| | Dyspnea | 14 | | 17 | | |
| | Dyspnea with Exertion | 9 | | 14 | | |
| | Chronic Cough | 13 | | 14 | | |
| | Atypical Chest Pain or Other | 12 | | 14 | | |
| ***Sequelae by SARS-CoV-2 Status*** | | | 166 | | 223 | |
| ***Sequelae per Patient by SARS-CoV-2 Status*** | | | 2.27 | | 2.23 | |
| ***Patients with Any Sequelae*** | | | 42 | | 48 | |
| ***Total Patients with Any Sequelae*** | | | | 90 | | |
| ***Total Sequelae*** | | | | 389 | | |
| ***Total Sequelae per Study Cohort Patient*** | | | | 2.25 | | |
| ***Sequelae per Patient with Any Sequelae (n = 90)*** | | | | 4.32 | | |

[*] Some sequelae counts have been combined to protect patient health information.

**Table S2. Quadratic Regression Models of Protein Quantities Dependent on Days from First Symptom**

| Uniprot Designation | Protein | Intercept | Intercept *p*-value | Linear Effect | Linear *p*-value | Curvature Effect | Curvature *p*-value | |
|---|---|---|---|---|---|---|---|---|
| | | | | | | | Unadjusted | FDR Adjusted |
| Q8IY21 | DDX60 | 10.829 | 3.34E-111 | 0.243 | 5.00E-09 | -0.009 | 5.04E-10 | 3.19E-06 |
| P10321 | HLA-C | 13.811 | 1.33E-153 | 0.151 | 1.73E-07 | -0.006 | 6.79E-09 | 1.98E-05 |
| P19525 | EIF2AK2 | 12.107 | 1.07E-151 | 0.127 | 9.81E-07 | -0.005 | 9.41E-09 | 1.98E-05 |
| P29728 | OAS2 | 12.572 | 1.96E-106 | 0.267 | 1.75E-07 | -0.011 | 1.29E-08 | 2.05E-05 |
| O95786 | RIGI | 12.662 | 2.23E-151 | 0.143 | 1.88E-07 | -0.006 | 2.06E-08 | 2.16E-05 |
| P20591 | MX1 | 12.825 | 3.38E-120 | 0.234 | 5.38E-08 | -0.009 | 2.28E-08 | 2.16E-05 |
| P04439 | HLA-A | 13.637 | 3.11E-149 | 0.144 | 1.82E-06 | -0.006 | 2.39E-08 | 2.16E-05 |
| P42224 | STAT1 | 13.742 | 1.23E-141 | 0.176 | 2.13E-07 | -0.007 | 3.27E-08 | 2.59E-05 |
| Q9Y6K5 | OAS3 | 12.345 | 1.16E-135 | 0.174 | 1.63E-07 | -0.007 | 3.84E-08 | 2.70E-05 |
| P29590 | PML | 12.607 | 3.37E-187 | 0.082 | 9.09E-07 | -0.003 | 5.43E-08 | 3.44E-05 |
| P01889 | HLA-B | 13.927 | 1.24E-146 | 0.149 | 2.92E-06 | -0.006 | 1.91E-07 | 1.10E-04 |
| P55265 | ADAR | 14.708 | 4.25E-192 | 0.076 | 2.58E-05 | -0.003 | 4.99E-07 | 2.63E-04 |
| Q8TDB6 | DTX3L | 11.471 | 9.60E-151 | 0.109 | 1.01E-05 | -0.005 | 5.85E-07 | 2.85E-04 |
| P80217 | IFI35 | 11.744 | 1.09E-154 | 0.100 | 2.66E-05 | -0.004 | 1.21E-06 | 5.45E-04 |
| Q8IXQ6 | PARP9 | 11.993 | 4.43E-149 | 0.110 | 2.88E-05 | -0.005 | 1.53E-06 | 6.46E-04 |
| P19474 | TRIM21 | 12.677 | 7.05E-162 | 0.092 | 7.54E-05 | -0.004 | 1.73E-06 | 6.84E-04 |
| Q03519 | TAP2 | 10.819 | 2.41E-139 | 0.123 | 6.32E-06 | -0.005 | 2.32E-06 | 8.63E-04 |
| P20592 | MX2 | 12.499 | 1.47E-113 | 0.192 | 2.16E-05 | -0.008 | 2.84E-06 | 9.99E-04 |
| Q9BYX4 | IFIH1 | 9.120 | 1.24E-78 | 0.242 | 1.23E-05 | -0.009 | 4.78E-06 | 1.59E-03 |
| Q13325 | IFIT5 | 11.051 | 1.37E-137 | 0.114 | 5.94E-05 | -0.005 | 6.13E-06 | 1.94E-03 |
| Q9Y3Z3 | SAMHD1 | 13.429 | 3.09E-176 | 0.081 | 7.10E-05 | -0.003 | 6.71E-06 | 2.02E-03 |
| Q63HN8 | RNF213 | 12.128 | 9.53E-176 | 0.068 | 2.21E-04 | -0.003 | 7.01E-06 | 2.02E-03 |
| Q08380 | LGALS3BP | 16.668 | 5.94E-157 | 0.130 | 7.07E-05 | -0.005 | 9.00E-06 | 2.48E-03 |
| A6NHR9 | SMCHD1 | 11.889 | 1.10E-162 | 0.084 | 9.76E-05 | -0.003 | 1.10E-05 | 2.90E-03 |
| P09914 | IFIT1 | 10.960 | 3.12E-88 | 0.208 | 2.51E-04 | -0.009 | 1.19E-05 | 3.02E-03 |
| Q8IYM9 | TRIM22 | 10.764 | 1.25E-104 | 0.157 | 3.35E-04 | -0.007 | 1.24E-05 | 3.02E-03 |
| P05161 | ISG15 | 11.816 | 6.85E-90 | 0.234 | 9.40E-05 | -0.009 | 2.46E-05 | 5.77E-03 |
| Q9H930 | SP140L | 10.667 | 1.24E-117 | 0.130 | 3.26E-04 | -0.006 | 2.91E-05 | 6.43E-03 |
| O15533 | TAPBP | 13.031 | 3.23E-161 | 0.092 | 1.43E-04 | -0.004 | 3.03E-05 | 6.43E-03 |
| P52630 | STAT2 | 11.591 | 6.81E-143 | 0.094 | 6.54E-04 | -0.004 | 3.06E-05 | 6.43E-03 |
| P14317 | HCLS1 | 12.391 | 4.92E-131 | 0.140 | 5.82E-05 | -0.005 | 3.15E-05 | 6.43E-03 |
| P42331 | ARHGAP25 | 11.279 | 1.69E-137 | 0.109 | 1.60E-04 | -0.004 | 3.45E-05 | 6.83E-03 |
| Q9H0J9 | PARP12 | 10.473 | 2.86E-113 | 0.135 | 3.43E-04 | -0.006 | 3.56E-05 | 6.83E-03 |
| Q9P2R3 | ANKFY1 | 12.509 | 7.75E-218 | 0.038 | 4.34E-04 | -0.002 | 3.70E-05 | 6.89E-03 |
| Q00978 | IRF9 | 9.696 | 3.52E-141 | 0.087 | 2.41E-04 | -0.004 | 3.96E-05 | 7.16E-03 |
| P28838 | LAP3 | 14.194 | 1.72E-190 | 0.067 | 1.44E-04 | -0.003 | 4.33E-05 | 7.61E-03 |
| Q13287 | NMI | 11.201 | 1.06E-131 | 0.113 | 2.96E-04 | -0.005 | 4.89E-05 | 8.36E-03 |
| P32455 | GBP1 | 12.200 | 4.17E-145 | 0.096 | 6.59E-04 | -0.004 | 5.29E-05 | 8.81E-03 |
| Q10589 | BST2 | 9.833 | 8.35E-93 | 0.168 | 4.12E-04 | -0.007 | 5.93E-05 | 9.27E-03 |
| Q16666 | IFI16 | 13.268 | 5.64E-178 | 0.066 | 7.14E-04 | -0.003 | 5.96E-05 | 9.27E-03 |
| Q01432 | AMPD3 | 11.654 | 1.41E-165 | 0.074 | 2.92E-04 | -0.003 | 6.00E-05 | 9.27E-03 |
| P32121 | ARRB2 | 12.713 | 2.29E-131 | 0.121 | 6.20E-04 | -0.005 | 6.34E-05 | 9.55E-03 |
| P28065 | PSMB9 | 12.421 | 3.60E-151 | 0.095 | 2.94E-04 | -0.004 | 6.62E-05 | 9.75E-03 |
| P19971 | TYMP | 13.749 | 2.24E-157 | 0.103 | 1.19E-04 | -0.004 | 6.93E-05 | 9.97E-03 |
| O15162 | PLSCR1 | 13.993 | 2.08E-125 | 0.166 | 9.59E-05 | -0.006 | 8.06E-05 | 1.13E-02 |
| A6NC98 | CCDC88B | 10.818 | 6.82E-117 | 0.125 | 7.21E-04 | -0.005 | 8.70E-05 | 1.20E-02 |
| P01893* | HLA-H* | 9.645 | 1.01E-77 | 0.209 | 3.67E-04 | -0.008 | 1.02E-04 | 1.37E-02 |
| Q86UX7 | FERMT3 | 15.397 | 9.72E-145 | 0.115 | 1.23E-03 | -0.005 | 1.10E-04 | 1.45E-02 |
| Q9UBW5 | BIN2 | 11.848 | 3.50E-126 | 0.119 | 7.70E-04 | -0.005 | 1.27E-04 | 1.64E-02 |
| Q13185 | CBX3 | 13.600 | 8.97E-169 | 0.073 | 1.16E-03 | -0.003 | 1.31E-04 | 1.66E-02 |
| Q92556 | ELMO1 | 11.754 | 1.51E-124 | 0.120 | 8.22E-04 | -0.005 | 1.39E-04 | 1.72E-02 |

**Table S2 (Continued). Quadratic Regression Models Protein Quantities Dependent on Days from First Symptom**

| Uniprot Designation | Protein | Intercept | Intercept *p*-value | Linear Effect | Linear *p*-value | Curvature Effect | Curvature *p*-value | |
|---|---|---|---|---|---|---|---|---|
| | | | | | | | Unadjusted | FDR Adjusted |
| Q96B97 | SH3KBP1 | 10.554 | 8.07E-109 | 0.147 | 2.87E-04 | -0.006 | 1.59E-04 | 1.91E-02 |
| P08670 | VIM | 16.773 | 1.67E-154 | 0.116 | 6.52E-04 | -0.005 | 1.60E-04 | 1.91E-02 |
| Q03518 | TAP1 | 12.832 | 9.11E-148 | 0.096 | 7.27E-04 | -0.004 | 1.65E-04 | 1.94E-02 |
| P09543 | CNP | 13.667 | 1.29E-198 | 0.053 | 4.48E-04 | -0.002 | 1.69E-04 | 1.95E-02 |
| Q9HD15 | SRA1 | 10.451 | 5.23E-150 | -0.081 | 3.20E-04 | 0.003 | 1.84E-04 | 2.06E-02 |
| P00973 | OAS1 | 12.224 | 7.30E-120 | 0.127 | 1.42E-03 | -0.005 | 1.85E-04 | 2.06E-02 |
| P41218 | MNDA | 15.786 | 3.63E-128 | 0.152 | 9.67E-04 | -0.006 | 1.90E-04 | 2.08E-02 |
| P08567 | PLEK | 12.733 | 7.27E-129 | 0.116 | 1.57E-03 | -0.005 | 2.01E-04 | 2.15E-02 |
| P29350 | PTPN6 | 14.056 | 6.59E-149 | 0.094 | 2.11E-03 | -0.004 | 2.05E-04 | 2.16E-02 |
| P21741 | MDK | 12.327 | 1.12E-128 | 0.135 | 1.63E-04 | -0.005 | 2.15E-04 | 2.20E-02 |
| Q9NUL5 | SHFL | 8.678 | 2.78E-101 | 0.113 | 2.25E-03 | -0.005 | 2.16E-04 | 2.20E-02 |
| P08133 | ANXA6 | 17.115 | 1.69E-138 | 0.137 | 1.53E-03 | -0.006 | 2.26E-04 | 2.27E-02 |
| Q460N5 | PARP14 | 11.127 | 9.62E-130 | 0.077 | 1.40E-02 | -0.004 | 2.43E-04 | 2.40E-02 |
| Q92608 | DOCK2 | 11.851 | 4.86E-135 | 0.101 | 1.27E-03 | -0.004 | 2.73E-04 | 2.66E-02 |
| Q9BYK8 | HELZ2 | 11.239 | 8.71E-143 | 0.078 | 3.25E-03 | -0.004 | 3.06E-04 | 2.89E-02 |
| P78540 | ARG2 | 11.082 | 1.91E-156 | -0.085 | 1.01E-04 | 0.003 | 3.06E-04 | 2.89E-02 |
| P26583 | HMGB2 | 14.907 | 1.03E-135 | 0.123 | 1.57E-03 | -0.005 | 3.16E-04 | 2.94E-02 |
| Q9BV57 | ADI1 | 12.116 | 1.80E-168 | -0.082 | 5.46E-05 | 0.003 | 3.46E-04 | 3.13E-02 |
| Q92843 | BCL2L2 | 10.704 | 1.13E-145 | -0.103 | 3.04E-05 | 0.003 | 3.46E-04 | 3.13E-02 |
| Q96RU3 | FNBP1 | 11.931 | 2.28E-162 | 0.069 | 1.48E-03 | -0.003 | 3.60E-04 | 3.21E-02 |
| A6NHX0 | CASTOR2 | 10.404 | 3.00E-149 | -0.081 | 3.77E-04 | 0.003 | 3.80E-04 | 3.31E-02 |
| Q9ULC5 | ACSL5 | 10.486 | 1.65E-135 | 0.097 | 4.30E-04 | -0.004 | 3.81E-04 | 3.31E-02 |
| P31146 | CORO1A | 14.995 | 8.17E-132 | 0.131 | 1.58E-03 | -0.005 | 4.03E-04 | 3.42E-02 |
| P13164 | IFITM1 | 11.145 | 2.44E-100 | 0.157 | 1.15E-03 | -0.006 | 4.08E-04 | 3.42E-02 |
| P61769 | B2M | 15.355 | 2.94E-145 | 0.102 | 3.59E-03 | -0.005 | 4.10E-04 | 3.42E-02 |
| O94804 | STK10 | 11.726 | 3.39E-173 | 0.059 | 1.35E-03 | -0.002 | 4.25E-04 | 3.48E-02 |
| P09913 | IFIT2 | 10.757 | 1.61E-109 | 0.114 | 5.14E-03 | -0.005 | 4.31E-04 | 3.48E-02 |
| O60234 | GMFG | 13.520 | 2.79E-137 | 0.099 | 4.10E-03 | -0.004 | 4.37E-04 | 3.48E-02 |
| Q9Y2Q0 | ATP8A1 | 11.816 | 9.92E-139 | 0.093 | 1.80E-03 | -0.004 | 4.44E-04 | 3.48E-02 |
| Q7Z2W4 | ZC3HAV1 | 12.336 | 2.06E-212 | 0.034 | 2.93E-03 | -0.001 | 4.46E-04 | 3.48E-02 |
| P13747 | HLA-E | 10.776 | 6.93E-137 | 0.084 | 2.33E-03 | -0.004 | 4.61E-04 | 3.55E-02 |
| O95466 | FMNL1 | 11.949 | 1.58E-139 | 0.083 | 4.93E-03 | -0.004 | 4.66E-04 | 3.55E-02 |
| P07305 | H1-0 | 15.715 | 4.21E-177 | 0.081 | 4.99E-04 | -0.003 | 4.72E-04 | 3.55E-02 |
| Q92835 | INPP5D | 11.413 | 1.82E-120 | 0.106 | 4.09E-03 | -0.005 | 5.03E-04 | 3.75E-02 |
| Q8TCU6 | PREX1 | 10.841 | 3.77E-130 | 0.089 | 3.49E-03 | -0.004 | 5.14E-04 | 3.78E-02 |
| Q8NHU6 | TDRD7 | 10.892 | 1.47E-174 | 0.047 | 4.53E-03 | -0.002 | 5.39E-04 | 3.89E-02 |
| O14727 | APAF1 | 11.701 | 1.45E-140 | 0.080 | 5.29E-03 | -0.004 | 5.42E-04 | 3.89E-02 |
| P17213 | BPI | 16.283 | 6.98E-119 | 0.161 | 2.78E-03 | -0.007 | 5.47E-04 | 3.89E-02 |
| Q92930 | RAB8B | 11.845 | 1.55E-128 | 0.094 | 6.15E-03 | -0.004 | 5.53E-04 | 3.89E-02 |
| Q14142 | TRIM14 | 9.822 | 1.44E-124 | 0.098 | 1.14E-03 | -0.004 | 5.89E-04 | 4.09E-02 |
| Q53GL7 | PARP10 | 10.343 | 2.44E-139 | 0.078 | 2.42E-03 | -0.003 | 5.95E-04 | 4.09E-02 |
| O75563 | SKAP2 | 11.669 | 4.17E-130 | 0.090 | 6.15E-03 | -0.004 | 6.49E-04 | 4.41E-02 |
| Q7L591 | DOK3 | 11.529 | 2.68E-114 | 0.113 | 5.51E-03 | -0.005 | 6.67E-04 | 4.41E-02 |
| Q9NUQ6 | SPATS2L | 10.388 | 6.81E-155 | 0.067 | 1.31E-03 | -0.003 | 6.68E-04 | 4.41E-02 |
| Q13303 | KCNAB2 | 10.548 | 1.93E-104 | 0.135 | 1.71E-03 | -0.005 | 6.68E-04 | 4.41E-02 |
| O95544 | NADK | 11.450 | 2.65E-103 | 0.130 | 6.01E-03 | -0.006 | 6.96E-04 | 4.54E-02 |
| P20160 | AZU1 | 18.550 | 1.19E-126 | 0.164 | 2.84E-03 | -0.007 | 7.03E-04 | 4.54E-02 |
| P98171 | ARHGAP4 | 12.295 | 2.54E-139 | 0.085 | 5.36E-03 | -0.004 | 7.35E-04 | 4.70E-02 |
| P55160 | NCKAP1L | 11.576 | 1.99E-140 | 0.079 | 5.30E-03 | -0.003 | 7.61E-04 | 4.81E-02 |
| P23526 | AHCY | 15.368 | 2.30E-224 | -0.043 | 3.11E-04 | 0.001 | 7.74E-04 | 4.85E-02 |

* P01893 *HLA-H* is annotated as a pseudogene at Uniprot.[1] We required identification of one unique peptide to report finding any specific protein. See **Methods** *Mass Spectrometry Data Analysis*. In this case, we identified the peptide YTCHVQHEGLPEPLTLR which

is unique to HLA-H. A recent work suggests that there are potential activities associated with HLA-H protein,[2] but further work is needed to confirm protein translation *in vivo,* perhaps involving targeted MS methods. **Abbreviation:** FDR false discovery rate.


**Table S3. Quadratic Regression Results for Soluble Human Saliva Proteins**

| **Biomarker** | | **Intercept** | **Intercept** | **Linear** | **Linear** | **Curvature** | **Curvature** |
|---|---|---|---|---|---|---|---|
| **Type** | **Protein** | | ***p*-value** | **Effect** | ***p*-value** | **Effect** | ***p*-value** |
| **Inflammatory Biomarker** | GMCSF | 5.601 | 2.77E-03 | -0.200 | 0.593 | 0.003 | 0.841 |
| | IL1β | 942.528 | 1.97E-03 | -17.916 | 0.769 | -0.310 | 0.888 |
| | IL6 | 334.217 | 7.12E-02 | -19.319 | 0.606 | 0.257 | 0.850 |
| | IL8 | 3349.728 | 2.57E-09 | 62.892 | 0.561 | -4.844 | 0.217 |
| | IL10 | 13.690 | 4.28E-03 | -0.995 | 0.301 | 0.019 | 0.584 |
| | IL12p70 | 1.329 | 6.72E-02 | 0.005 | 0.971 | -0.002 | 0.775 |
| | IP10 | 631.843 | 9.32E-03 | 27.344 | 0.576 | -1.788 | 0.313 |
| | TNFα | 217.100 | 1.77E-01 | -19.376 | 0.552 | 0.420 | 0.721 |
| **Type I Interferon** | IFN$\alpha_2$ | 15.992 | 1.34E-01 | -1.503 | 0.487 | 0.033 | 0.670 |
| | IFNβ | 6.613 | 8.22E-03 | 0.125 | 0.804 | -0.011 | 0.558 |
| **Type II Interferon** | IFNγ | 19.539 | 2.63E-02 | -1.277 | 0.472 | 0.020 | 0.753 |
| **Type III Interferon** | IFN$\lambda_1$ | 44.225 | 3.96E-03 | -3.184 | 0.301 | 0.072 | 0.517 |
| | IFN$\lambda_2$ | 24.406 | 1.66E-04 | -0.381 | 0.767 | -0.006 | 0.891 |

**Table S4. Viral Detections**

| Virus Detection | Patients Affected (fraction)[‡] |
|---|---|
| *Adenovirus* species[*] | < 5[*] |
| *Human Metapneumovirus*[*] | < 5[*] |
| Rhinoviruses A, B or C or other *Enterovirus* | 13 (0.0751) |
| Non-SARS-CoV-2 coronaviruses | 7 (0.0405) |
| Influenza, any type[*] | < 5[*] |
| *Parainfluenza* Types 1-4[*] | < 5[*] |
| Non SARS-CoV-2 Detections by Respiratory Viral Panels[†] | 19 (0.11) |
| Viral Detections by RNA-Seq | 54 (0.403)[‡] |
| Multiple Viral Detections[§] | 14 (0.0809) |
| No Viral Detections by any method[†] | 60 (0.347) |
| All SARS-CoV-2 Detections | 100 (0.578) |

[*]Precise number not provided due to patient privacy concerns. [†] Respiratory Viral Panels gradually increased in availability and in numbers of viruses detected throughout the study, complicating interpretation of this result. [§] Includes 6 patients with SARS-CoV-2 infections. [‡] Fraction is based on the number of samples that underwent RNA-Seq, 134.

**PCR and RNA-Seq Detections of SARS-CoV-2 from Simultaneous Samples**

| | | RNA-Seq | | |
|---|---|---|---|---|
| | | **Negative** | **Positive** | **Totals** |
| **PCR** | **Negative** | 31 | 10 | 41 |
| | **Positive** | 8 | 19 | 27 |
| | **Totals** | 39 | 29 | n = 68 |

| RNA-Seq as Reference Test | |
|---|---|
| **PCR False Negative Rate** | PCR False Negative/PCR All Negatives = 10/(31+10)= 0.244 |
| **PCR False Positive Rate** | PCR False Positive/PCR All Positives = 8/(8+19) = 0.296 |

| PCR as Reference Test | |
|---|---|
| **RNA-Seq False Negative Rate** | RNA-Seq False Negative/RNA-Seq All Negative = 8/(31+8) = 0.205 |
| **RNA-Seq False Positive Rate** | RNA-Seq False Positive/RNA-Seq All Positive = 10/(10+19) = 0.345 |

**Table S5. Principal Components for Intrinsic and Comorbidity Variables.**[*]

| Variable | Principal Components | | |
|---|---|---|---|
| | **MedPC1** | **MedPC2** | **MedPC3** |
| Age | 0.296 | -0.260 | -0.243 |
| Sex | 0.072 | 0.171 | 0.146 |
| CNS Disease | 0.147 | 0.210 | 0.298 |
| Mental Health Disease | 0.121 | 0.258 | 0.313 |
| Hypertension | 0.216 | -0.220 | 0.233 |
| Heart Disease | 0.280 | -0.274 | 0.031 |
| Lung Disease | 0.190 | -0.172 | 0.097 |
| Bleeding Diseases | 0.193 | 0.024 | 0.024 |
| Gastrointestinal Disease | 0.289 | 0.150 | 0.099 |
| *Diabetes mellitus* | 0.161 | -0.212 | 0.335 |
| Obesity | 0.160 | -0.190 | 0.387 |
| Endocrine Diseases | 0.202 | 0.064 | -0.191 |
| Rheumatologic Diseases | 0.299 | 0.015 | 0.025 |
| Chronic Infections | 0.009 | -0.026 | -0.357 |
| Cancer | 0.225 | 0.054 | -0.345 |
| Kidney Disease | 0.237 | 0.102 | -0.068 |
| Prior Year Clinic Visits | 0.322 | 0.174 | 0.053 |
| Prior Year ED Visits | 0.259 | 0.270 | -0.159 |
| Prior Year Hospital Visits | 0.329 | 0.230 | -0.211 |
| Hypoxemia in ED | 0.061 | -0.459 | -0.031 |
| Admission from ED | 0.132 | -0.388 | -0.181 |
| Standard deviation | 2.166 | 1.450 | 1.268 |
| Proportion of Variance | 0.224 | 0.100 | 0.077 |
| Cumulative Proportion | 0.224 | 0.324 | 0.400 |

[*] Raw values were centered and scaled during computation. All components in MedPC1 are positive indicating that high MedPC1 represents the presence of some or all past and concurrent comorbidities. Mixed signs make interpretation of other principal components difficult if not impossible.

**Table S6. Principal Components for 44 Proteins Clinically Relevant for Viral Syndromes[*]**

| Uniprot Designation | Protein | Principal Components | | |
|---|---|---|---|---|
| | | QuadPC1 | QuadPC2 | QuadPC3 |
| Q8IY21 | DDX60 | 0.173 | 0.118 | -0.056 |
| P10321 | HLA-C | 0.144 | -0.168 | -0.009 |
| P19525 | EIF2AK2 | 0.181 | 0.037 | 0.017 |
| P29728 | OAS2 | 0.180 | 0.126 | -0.125 |
| O95786 | RIGI | 0.181 | 0.034 | -0.070 |
| P20591 | MX1 | 0.165 | 0.168 | -0.218 |
| P04439 | HLA-A | 0.155 | -0.191 | 0.054 |
| P42224 | STAT1 | 0.169 | 0.184 | -0.066 |
| Q9Y6K5 | OAS3 | 0.173 | 0.077 | -0.185 |
| P29590 | PML | 0.170 | -0.122 | 0.058 |
| P01889 | HLA-B | 0.133 | -0.213 | -0.125 |
| P55265 | ADAR | 0.144 | -0.139 | -0.019 |
| Q8TDB6 | DTX3L | 0.167 | 0.028 | 0.075 |
| P80217 | IFI35 | 0.149 | -0.027 | 0.256 |
| Q8IXQ6 | PARP9 | 0.176 | 0.104 | 0.160 |
| P19474 | TRIM21 | 0.165 | -0.150 | 0.053 |
| Q03519 | TAP2 | 0.148 | 0.043 | 0.119 |
| P20592 | MX2 | 0.158 | 0.198 | 0.049 |
| Q9BYX4 | IFIH1 | 0.163 | 0.080 | -0.097 |
| Q13325 | IFIT5 | 0.154 | 0.133 | 0.142 |
| Q9Y3Z3 | SAMHD1 | 0.162 | -0.009 | 0.022 |
| Q63HN8 | RNF213 | 0.180 | -0.053 | 0.208 |
| Q08380 | LGALS3BP | 0.133 | 0.025 | -0.239 |
| A6NHR9 | SMCHD1 | 0.093 | -0.323 | 0.113 |
| P09914 | IFIT1 | 0.160 | 0.089 | -0.229 |
| Q8IYM9 | TRIM22 | 0.163 | -0.072 | -0.077 |
| P05161 | ISG15 | 0.153 | 0.175 | -0.263 |
| Q9H930 | SP140L | 0.162 | -0.032 | 0.045 |
| O15533 | TAPBP | 0.133 | 0.009 | 0.249 |
| P52630 | STAT2 | 0.172 | 0.000 | 0.117 |
| P14317 | HCLS1 | 0.059 | -0.227 | -0.336 |
| P42331 | ARHGAP25 | 0.083 | -0.289 | 0.010 |
| Q9H0J9 | PARP12 | 0.167 | 0.041 | 0.111 |
| Q9P2R3 | ANKFY1 | 0.137 | 0.052 | 0.322 |
| Q00978 | IRF9 | 0.142 | 0.097 | -0.112 |
| P28838 | LAP3 | 0.141 | 0.209 | 0.210 |
| Q13287 | NMI | 0.143 | -0.125 | -0.004 |
| P32455 | GBP1 | 0.156 | 0.045 | 0.191 |
| Q10589 | BST2 | 0.162 | 0.099 | -0.182 |
| Q16666 | IFI16 | 0.154 | -0.176 | -0.086 |
| Q01432 | AMPD3 | 0.093 | -0.249 | -0.029 |
| P32121 | ARRB2 | 0.091 | -0.339 | -0.136 |
| P28065 | PSMB9 | 0.122 | -0.186 | -0.001 |
| P19971 | TYMP | 0.105 | 0.199 | -0.148 |
| | Standard deviation | 5.048 | 2.348 | 1.217 |
| | Proportion of Variance | 0.579 | 0.125 | 0.034 |
| | Cumulative Proportion | 0.579 | 0.704 | 0.738 |

[*] Raw values were centered and scaled prior to computation. Protein ordered by $p_{FDR}$ of curvature term in quadratic regression of measurements by days from first symptoms. All values of QuadPC1 are positive indicating that an increase in QuadPC1 indicates an increase in all the included proteins. The 44 proteins were included because they had $p_{FDR} < 0.01$ in quadratic regression against days from first viral syndrome symptom.

**Table S7. Protein Incidence Ratios (IR) Associated with Decreased Subsequent Neurocognitive Sequelae***

| Uniprot Designation | Gene | $Log_2$ Protein Range | SARS-CoV-2 | | | Protein | | | SARS-CoV-2-Protein Interaction | | |
|---|---|---|---|---|---|---|---|---|---|---|---|
| | | | IR | 95% CI | *p* | IR | 95% CI | *p* | IR | 95% CI | *p* |
| Q00978 | IRF9 | 9.49-12.13 | 1.05 | 0.63-1.75 | 0.86 | 1.91 | 1.000-3.62 | 0.05 | 0.251 | 0.109-0.576 | 0.001 |
| P28838 | LAP3 | 13.09-15.99 | 1.20 | 0.69-2.10 | 0.51 | 1.37 | 0.498-3.76 | 0.54 | 0.265 | 0.078-0.900 | 0.033 |
| P29590 | PML | 11.67-14.16 | 1.18 | 0.70-1.99 | 0.53 | 1.29 | 0.546-3.04 | 0.56 | 0.304 | 0.099-0.929 | 0.037 |
| Q9Y6K5 | OAS3 | 11.16-15.49 | 0.93 | 0.53-1.65 | 0.81 | 1.76 | 0.974-3.18 | 0.06 | 0.320 | 0.161-0.636 | 0.001 |
| P80217 | IFI35 | 10.39-13.90 | 1.12 | 0.68-1.86 | 0.65 | 1.41 | 0.844-2.37 | 0.19 | 0.320 | 0.153-0.668 | 0.002 |
| P42224 | STAT1 | 12.24-17.07 | 0.98 | 0.56-1.72 | 0.95 | 1.52 | 0.903-2.57 | 0.11 | 0.335 | 0.178-0.631 | <0.001 |
| P55265 | ADAR | 13.65-17.54 | 1.26 | 0.75-2.12 | 0.39 | 0.85 | 0.437-1.67 | 0.64 | 0.340 | 0.123-0.937 | 0.037 |
| P19971 | TYMP | 12.34-15.96 | 1.13 | 0.69-1.86 | 0.62 | 1.39 | 0.853-2.26 | 0.19 | 0.344 | 0.183-0.645 | <0.001 |
| Q8TDB6 | DTX3L | 9.91-14.16 | 1.13 | 0.66-1.96 | 0.65 | 1.48 | 0.734-2.97 | 0.27 | 0.351 | 0.157-0.785 | 0.011 |
| P20591 | MX1 | 11.24-16.74 | 0.94 | 0.52-1.71 | 0.85 | 1.48 | 0.939-2.33 | 0.09 | 0.351 | 0.202-0.610 | <0.001 |
| P19525 | EIF2AK2 | 10.81-14.70 | 1.17 | 0.67-2.02 | 0.58 | 1.18 | 0.660-2.11 | 0.58 | 0.394 | 0.189-0.820 | 0.013 |
| P19474 | TRIM21 | 11.42-14.72 | 1.15 | 0.69-1.91 | 0.59 | 1.12 | 0.654-1.92 | 0.68 | 0.430 | 0.209-0.882 | 0.021 |
| Q8IXQ6 | PARP9 | 9.81-14.36 | 1.22 | 0.69-2.16 | 0.49 | 1.20 | 0.637-2.24 | 0.58 | 0.442 | 0.210-0.930 | 0.031 |
| Q8IY21 | DDX60 | 9.79-14.66 | 1.06 | 0.62-1.83 | 0.83 | 1.32 | 0.879-1.98 | 0.18 | 0.452 | 0.278-0.735 | 0.001 |
| O15533 | TAPBP | 11.66-15.72 | 1.17 | 0.70-1.97 | 0.54 | 1.49 | 0.852-2.62 | 0.16 | 0.455 | 0.212-0.975 | 0.043 |
| Q08380 | LGALS3BP | 15.17-20.28 | 1.16 | 0.69-1.95 | 0.59 | 1.37 | 0.921-2.03 | 0.12 | 0.459 | 0.271-0.778 | 0.004 |
| Q10589 | BST2 | 9.28-14.41 | 0.96 | 0.54-1.71 | 0.89 | 1.59 | 1.050-2.40 | 0.03 | 0.460 | 0.284-0.747 | 0.002 |
| Q9H930 | SP140L | 9.07-13.24 | 1.13 | 0.68-1.90 | 0.63 | 1.40 | 0.888-2.19 | 0.15 | 0.490 | 0.288-0.836 | 0.009 |
| Q03519 | TAP2 | 9.36-14.36 | 1.15 | 0.68-1.93 | 0.60 | 1.54 | 0.893-2.64 | 0.12 | 0.491 | 0.245-0.981 | 0.044 |
| P29728 | OAS2 | 10.39-17.08 | 1.15 | 0.60-2.20 | 0.67 | 1.17 | 0.784-1.75 | 0.44 | 0.601 | 0.380-0.951 | 0.030 |
| P05161 | ISG15 | 9.51-17.07 | 1.12 | 0.62-2.03 | 0.70 | 1.24 | 0.904-1.69 | 0.18 | 0.606 | 0.419-0.876 | 0.008 |
| Q8IYM9 | TRIM22 | 9.13-14.8 | 1.24 | 0.73-2.10 | 0.43 | 1.07 | 0.785-1.45 | 0.68 | 0.636 | 0.431-0.938 | 0.022 |
| P09914 | IFIT1 | 9.50-15.04 | 1.42 | 0.80-2.55 | 0.23 | 1.01 | 0.757-1.34 | 0.96 | 0.690 | 0.488-0.978 | 0.037 |

*Results shown for models adjusted for Mental Health and CNS diseases. Proteins are listed in the order of decreasing impact of SARS-CoV-2-Protein Interaction Incidence Ratio. Because all protein values underwent $log_2$ transformation prior to modeling, a one unit increase in $log_2$-transformed protein equals a doubling of detected amount. For each protein, the impact of a doubling of protein is described by a fractional decrease in neurocognitive sequelae equal to the Interaction Incidence Ratio in SARS-CoV-2 patients. In interaction models, the incidence ratios for the linear interactors should be ignored. For example, for a doubling of IRF9 measured, there is an estimated associated decrease in subsequent sequelae by a factor of 0.25, or a 75% reduction in sequelae count. In patients without SARS-CoV-2, there is no change in neurocognitive sequelae associated with a change in IRF9. For IFIT1, the estimated associated decrease in neurocognitive sequelae is 31% because of the estimated interaction ratio of 0.69. **Abbreviation**: CI confidence interval.

**Table S8. Proteins Associated with Decreased Subsequent General Sequelae**

| Uniprot Designation | Gene | $Log_2$ Protein Range | SARS-CoV-2 | | | Protein | | |
|---|---|---|---|---|---|---|---|---|
| | | | Incidence Ratio | 95% CI | *p* | Incidence Ratio | 95% CI | *p* |
| P55265 | ADAR | 13.65 to 17.54 | 1.78 | 1.03 to 3.071 | 0.039 | 0.41 | 0.244 to 0.694 | 0.001 |
| Q9P2R3 | ANKFY1 | 11.78 to 13.68 | 1.54 | 0.897 to 2.661 | 0.117 | 0.41 | 0.185 to 0.923 | 0.031 |
| Q63HN8 | RNF213 | 10.96 to 14.1 | 1.72 | 0.991 to 2.978 | 0.054 | 0.51 | 0.318 to 0.821 | 0.006 |
| P28838 | LAP3 | 13.09 to 15.99 | 1.73 | 0.977 to 3.051 | 0.060 | 0.54 | 0.323 to 0.897 | 0.017 |
| P19474 | TRIM21 | 11.42 to 14.72 | 1.65 | 0.945 to 2.87 | 0.078 | 0.61 | 0.422 to 0.888 | 0.010 |
| P19525 | EIF2AK2 | 10.81 to 14.7 | 1.83 | 1.026 to 3.248 | 0.041 | 0.64 | 0.453 to 0.901 | 0.011 |
| O95786 | RIGI | 11.69 to 15.68 | 1.89 | 1.038 to 3.426 | 0.037 | 0.65 | 0.462 to 0.921 | 0.015 |
| P52630 | STAT2 | 9.87 to 13.95 | 1.58 | 0.904 to 2.757 | 0.108 | 0.71 | 0.515 to 0.97 | 0.032 |
| Q13325 | IFIT5 | 8.98 to 13.79 | 1.67 | 0.935 to 2.995 | 0.083 | 0.72 | 0.522 to 0.996 | 0.048 |
| P20591 | MX1 | 11.24 to 16.74 | 1.95 | 1.039 to 3.662 | 0.038 | 0.74 | 0.584 to 0.938 | 0.013 |
| Q8IYM9 | TRIM22 | 9.13 to 14.8 | 1.66 | 0.957 to 2.866 | 0.071 | 0.74 | 0.612 to 0.904 | 0.003 |
| P42224 | STAT1 | 12.24 to 17.07 | 1.69 | 0.931 to 3.059 | 0.084 | 0.75 | 0.568 to 0.993 | 0.044 |
| Q9H930 | SP140L | 9.07 to 13.24 | 1.52 | 0.885 to 2.614 | 0.129 | 0.76 | 0.603 to 0.962 | 0.022 |
| Q8IY21 | DDX60 | 9.79 to 14.66 | 1.74 | 0.977 to 3.100 | 0.060 | 0.77 | 0.618 to 0.949 | 0.015 |
| P01889 | HLA-B | 10.57 to 16.58 | 1.46 | 0.853 to 2.497 | 0.167 | 0.77 | 0.595 to 0.997 | 0.048 |
| P29728 | OAS2 | 10.39 to 17.08 | 2.00 | 1.074 to 3.709 | 0.029 | 0.78 | 0.647 to 0.95 | 0.013 |
| Q10589 | BST2 | 9.28 to 14.41 | 1.76 | 0.963 to 3.206 | 0.066 | 0.80 | 0.654 to 0.985 | 0.036 |
| P05161 | ISG15 | 9.51 to 17.07 | 1.83 | 0.991 to 3.369 | 0.054 | 0.83 | 0.704 to 0.975 | 0.024 |
| P09914 | IFIT1 | 9.5 to 15.04 | 1.70 | 0.937 to 3.071 | 0.081 | 0.83 | 0.704 to 0.981 | 0.028 |

*Results shown reflect adjustments for age and past medical histories of heart disease and obesity. There were no significant interactions between proteins and SARS-CoV-2. Presence of SARS-CoV-2 increased the average number of General Sequelae by a factor equal to the Incidence Ratio shown while for every unit increase in $log_2$ transformed protein amount, the count of General Sequelae is reduced by a factor equal to the Incidence Ratio. For example, in the case of ADAR, each unit increase in $log_2$-transformed ADAR, was associated with a reduction in the average General Sequelae count by a factor of 0.41 or a 59% decrease while a 2 unit increase in $log_2$ transformed ADAR was associated with a reduction in sequelae by a factor of 0.41 × 0.41 = 0.1681, or a reduction of ~83%. The Incidence Ratio for SARS-CoV-2 ranged from 1.46 to 2.00 with a mean of 1.71 suggesting that General Sequelae counts were associated with an increase by a factor of 1.71 or 71% with COVID19 infection, independent of any protein spectral intensity. **Abbreviation**: CI confidence interval.

**Table S9. Reactome Pathways**[*]

| Pathway Identifier | Pathway Name | Found Proteins | Total Pathway Proteins | *p*-value | FDR adjusted *p*-value |
|---|---|---|---|---|---|
| ***Pathways Involving Proteins Associated with Neurocognitive Sequelae*** | | | | | |
| R-HSA-75064 | mRNA Editing: A to I Conversion | 1 | 2 | 4.43E-03 | 2.05E-02 |
| R-HSA-77042 | Formation of editosomes by ADAR proteins | 1 | 2 | 4.43E-03 | 2.05E-02 |
| R-HSA-169131 | Inhibition of PKR | 1 | 2 | 4.43E-03 | 2.05E-02 |
| R-HSA-75102 | C6 deamination of adenosine | 1 | 2 | 4.43E-03 | 2.05E-02 |
| R-HSA-909733 | Interferon alpha/beta signaling | 13 | 80 | 1.11E-16 | 8.44E-15 |
| R-HSA-877300 | Interferon gamma signaling | 7 | 98 | 1.71E-09 | 6.49E-08 |
| R-HSA-1169408 | ISG15 antiviral mechanism | 5 | 80 | 8.84E-07 | 1.86E-05 |
| R-HSA-913531 | Interferon Signaling | 16 | 361 | 1.11E-16 | 8.44E-15 |
| R-HSA-1169410 | Antimicrobial mechanism of IFN-stimulated genes | 8 | 237 | 3.33E-08 | 9.99E-07 |
| R-HSA-1280215 | Cytokine Signaling in Immune system | 16 | 861 | 2.45E-12 | 1.23E-10 |
| R-HSA-9694516 | SARS-CoV-2 Infection | 5 | 320 | 6.29E-04 | 6.29E-03 |
| R-HSA-9679506 | SARS-CoV Infections | 7 | 517 | 1.05E-04 | 1.58E-03 |
| R-HSA-9824446 | Viral Infection Pathways | 12 | 1285 | 7.32E-06 | 1.39E-04 |
| R-HSA-168256 | Immune System | 19 | 2526 | 4.17E-08 | 1.04E-06 |
| R-HSA-5663205 | Infectious disease | 12 | 1732 | 1.49E-04 | 1.94E-03 |
| ***Pathways Involving Proteins Associated with General Sequelae*** | | | | | |
| R-HSA-75064 | mRNA Editing: A to I Conversion | 1 | 2 | 3.45E-03 | 1.03E-02 |
| R-HSA-77042 | Formation of editosomes by ADAR proteins | 1 | 2 | 3.45E-03 | 1.03E-02 |
| R-HSA-169131 | Inhibition of PKR | 1 | 2 | 3.45E-03 | 1.03E-02 |
| R-HSA-75102 | C6 deamination of adenosine | 1 | 2 | 3.45E-03 | 1.03E-02 |
| R-HSA-909733 | Interferon alpha/beta signaling | 13 | 80 | 1.11E-16 | 7.66E-15 |
| R-HSA-1169408 | ISG15 antiviral mechanism | 6 | 80 | 4.05E-09 | 9.32E-08 |
| R-HSA-877300 | Interferon gamma signaling | 5 | 98 | 6.22E-07 | 8.09E-06 |
| R-HSA-9705671 | SARS-CoV-2 activates/modulates innate and adaptive immune responses | 7 | 144 | 3.29E-09 | 8.89E-08 |
| R-HSA-9833110 | RSV-host interactions | 5 | 108 | 1.00E-06 | 1.10E-05 |
| R-HSA-913531 | Interferon Signaling | 16 | 361 | 1.11E-16 | 7.66E-15 |
| R-HSA-1169410 | Antimicrobial mechanism of IFN-stimulated genes | 9 | 237 | 9.68E-11 | 3.29E-09 |
| R-HSA-9820952 | Respiratory Syncytial Virus Infection Pathway | 5 | 133 | 2.76E-06 | 2.48E-05 |
| R-HSA-9705683 | SARS-CoV-2-host interactions | 7 | 221 | 6.10E-08 | 1.04E-06 |
| R-HSA-9694516 | SARS-CoV-2 Infection | 7 | 320 | 7.36E-07 | 8.83E-06 |
| R-HSA-1280215 | Cytokine Signaling in Immune system | 16 | 861 | 5.77E-15 | 2.66E-13 |
| R-HSA-9679506 | SARS-CoV Infections | 9 | 517 | 8.39E-08 | 1.26E-06 |
| R-HSA-9824446 | Viral Infection Pathways | 11 | 1285 | 2.36E-06 | 2.36E-05 |
| R-HSA-5663205 | Infectious disease | 12 | 1732 | 5.82E-06 | 5.24E-05 |
| R-HSA-168256 | Immune System | 17 | 2526 | 6.29E-09 | 1.19E-07 |
| R-HSA-1643685 | Disease | 12 | 2580 | 3.50E-04 | 2.10E-03 |

[*]Selected pathways with an FDR adjusted *p*-value $< 0.05$ that involved at least 5 of the proteins found in saliva that predicted Neurocognitive or General Sequelae OR the saliva proteins represented more than 30% of entire small pathways. Pathways are arranged in decreasing order of the percentage of clinically relevant saliva proteins found among the total proteins in each Reactome pathway.
